# High Respiratory Syncytial Virus Burden in Children Under 3 Years of Age Across All Care Levels in England

**DOI:** 10.1101/2025.07.14.25331490

**Authors:** Emma C. Carter, Helen Hill, Carla Solórzano, Lauren Kerruish, Lauren McLellan, James Dodd, Adam B. Smith, Andria Joseph, Damian Lewis, Fred Fyles, Simon B. Drysdale, Patricia Gonzalez-Dias, Gregory S.J. Duncan, Kelly Davies, Paula Saunderson, Mathieu Bangert, Rolf Kramer, Natalya Vassilouthis, Maia Lesosky, Shrouk Messahel, Caroline Burchett, Stephen Brearey, Jolanta Bernatoniene, Hayley King, Sudeshna Bhowmik, James Perry, Rebecca Sinfield, Pat Mottram, Raqib Huq, Paul McNamara, David Lewis, Nadja Van Ginneken, Daniela M. Ferreira, Andrea M. Collins, the STOP RSV investigators

## Abstract

Recent evidence has shown a substantial RSV burden in healthy term-born children, but there is less data quantifying the relative burden compared to those with co-morbidity or prematurity. There is also limited primary care and emergency department (ED) data due to the lack of routine pathogen detection. These data are important for the development of RSV immunisation strategies.

A prospective surveillance study of children < 3 years old, presenting with lower respiratory infection and a sub-study in primary care of upper respiratory infection was conducted. The primary endpoint was RSV prevalence by healthcare setting. Secondary endpoints included proportion of hospitalisations, level of ventilatory support and admission to higher levels of care. An economic analysis assessing the costs associated with healthcare utilisation was also conducted.

The primary analysis revealed RSV prevalence was 50.4% in children admitted to hospital, 36.3% in ED discharges, 36.5% in primary care and 12.8% in primary care children with upper respiratory infection. Healthy term-born children accounted for 73.3% of medically-attended RSV cases and 70.1% of hospitalisations. Risk factors for severe disease included any level of prematurity, age < 3 months and congenital cardiac disease. RSV positive cases incurred a higher mean cost than RSV negative cases (mean = £1,706; incremental = £683) per illness.

The findings revealed a substantial burden associated with RSV. Even moderate prematurity was a risk factor for severe disease, and these children may not benefit from the full maternal immune response to vaccination and would not be eligible for Nirsevimab under UK guidance, we therefore recommend broadening eligibility to include these children.

**What is already known on this topic:** Recent cohort studies have revealed a substantial healthcare and economic RSV burden from previously healthy term-born children which has supported the implementation of a maternal RSV vaccination programme in the UK, providing protection for infants from birth up to 6 months old.

**What this study adds:** We have shown that children born prematurely, including moderately preterm children, are at increased risk of severe RSV disease. These children may not benefit from full protection by maternal RSV vaccination, and may not be eligible for Nirsevimab, the long-acting monoclonal antibody, which is currently reserved for only the most high-risk children in the UK.

**How this study might affect research, practice or policy:** This data supports broadening eligibility for Nirsevimab, to include preterm children, who were born before full maternal protection from RSV vaccination may have developed, and to infants born to mothers not receiving maternal vaccination.

## Introduction

Respiratory syncytial virus (RSV) is the most common respiratory infection in young children, causing a global epidemic of lower respiratory tract infections (LRTI), with over 33 million episodes estimated in <5-year-olds annually (1, 2, 3, 4). Furthermore, 5.5% of RSV-associated hospitalisations of healthy term babies require paediatric intensive care unit (PICU) admission (5).

Given this substantial burden, developing preventive therapies has been a priority. The first long-acting monoclonal antibody for infants - Nirsevimab - was licensed in the United Kingdom (UK) in November 2022(6). The first bivalent RSV vaccine (Pfizer RSVpreF) was licensed by the Food and Drugs Administration (FDA) for pregnant women in August 2023(7), by the European Medicines Agency in September 2023(8), and by the UK Medicines and Healthcare products Regulatory Approval (MHRA) in November 2023(9). Based on recommendations from UK Joint Committee on Vaccination and Immunisation (JCVI), vaccination for pregnant women over 28 weeks of gestation commenced in September 2024 (10). Maternal vaccination offers protection to infants for the first 6 months of life and is recommended at week 28. However, children who are born premature, may not fully benefit from the development of maternal immunity and would not be eligible for Nirsevimab, reserved for the most high-risk infants under current UKHSA guidance.

A recent European birth cohort study has been published on the burden of RSV in healthy term born infants, demonstrating a high burden of RSV in this cohort(5, 11). Various risk factors have been reported for severe RSV disease (12, 13), but there is less data available comparing the relative burden on healthcare settings from children with co-morbidity and prematurity and those without. Furthermore, there is limited UK data on the burden in primary care and emergency department discharges, as pathogen detection is not routinely conducted.

This prospective surveillance study aimed to estimate the prevalence of laboratory-confirmed RSV in children (<3 years) presenting to primary, secondary and tertiary care with RTI symptoms across all seasons. Moreover, we aimed to investigate the relative healthcare burden from healthy term-born children and those with co-morbidity or prematurity to inform developing immunisation strategies.

## 1. Materials and Methods

This study was registered with ISRCTN 08/08/2022 ISRCTN41075797. Full Research Ethics Committee (REC) approval (21/WS/0142, 02 Dec 2021) and Health Research Authority (HRA) (304483, 21^st^ December 2021) approval was granted prior to study commencement.

### Study Participants

The study was implemented in Merseyside, Cheshire (Northern England) and Bristol (Southern England), encompassing 11 primary care (general practice) sites, 5 walk-in centres (WIC), 2 secondary care hospital sites, and 2 tertiary care hospital sites. The study period spanned from December 9, 2021, to March 31, 2023. Children under three years presenting to primary, secondary and tertiary settings with lower respiratory tract infection (LRTI) symptoms were included (Supplementary Table 1). A sub-study (minimum n=180) was conducted in primary care to assess the prevalence of RSV-attributable upper respiratory tract infection (URTI) in children presenting with RTI symptoms without LRTI features.

Participants could be re-enrolled if they presented again after more than 30 days, indicating a new illness episode.

### Study Design

#### Objective

The primary objective of this study was to estimate the prevalence of laboratory confirmed RSV infection in children < 3 years of age presenting to primary, secondary and tertiary healthcare settings with respiratory tract infection symptoms.

#### Sample Size Calculation

The sample size target of 1,800 was calculated to achieve 90% power with 0.028 precision using a 2-sided exact test, with statistical significance 0.05, for the primary outcome, based on an estimated RSV positivity rate of 0.15 of nasal swabs.

### Methods

The protocol has been described previously (14). In summary, children meeting inclusion criteria (supplementary table 1) were recruited to the study and nasal swabs were conducted on the day of recruitment and processed at local NHS-associated laboratories by multiplex panel (supplementary table 2). Written consent was taken on recruitment, or in some cases verbal consent was taken for the nasal swab and full written consent taken as soon as possible. Results were made available to treating clinicians. Recruitment took place in primary care, secondary care and tertiary care from December 2021 to March 2023 and in walk-in-centres from December 2022 to March 2023.

#### Definitions

Hospitalisation was defined as an overnight admission to a ward. Prolonged respiratory symptoms were defined as ongoing respiratory symptoms for at least 14 days. A significant co-morbidity was defined according to expert consensus with paediatric specialists (included in the author publication list), and previously published literature on risk factors for severe RSV infection (23) (Table 3). Prematurity was categorised into extremely preterm (<28 weeks); very preterm (28 weeks - <32 weeks); moderate to late preterm (32 - <37 weeks).

Severe outcomes were defined as hospitalisation, admission to HDU or PICU, requirement for respiratory support.

### Outcomes and statistical analyses

#### Primary Endpoint

The primary endpoint of the study was the prevalence of RSV infection in primary, secondary and tertiary healthcare settings. Respiratory nasal swabs were collected on the day of recruitment and processed at local NHS-affiliated laboratories (Supplementary Table 2). The prevalence of RSV was reported as the proportion of nasal swabs positive for RSV.

#### Secondary Endpoints

To analyse participant outcomes, risk factors for severe disease, burden of disease, and prevalence of co-infection the following analyses were performed. Baseline data were obtained from medical records and parents/guardians. After 28 days, medical records were reviewed retrospectively to gather outcome data. Parents/guardians completed electronic questionnaires on days 14 and 28 to assess the duration of symptoms, additional healthcare contacts, and the impact of illness on work and caregiving responsibilities. Participants were classified into distinct levels of healthcare for analysis:

- Primary care (General Practice)
- Walk-in-centre (WIC)
- Emergency department (ED) + discharge - attended ED but were not admitted
- ED + admission - attended ED and were admitted
- Secondary and Tertiary Hospital admission (direct admission which could include a referral from primary care/WIC or an inter-hospital transfer)

Frequencies and proportions were used to summarise most data. For comparison of number of healthcare visits between RSV positive and negative participants, Chi-squared test was performed. The prevalence of viral co-infection was reported as the proportion of nasal swabs with detection of 2 or more viruses by PCR. Duration of symptoms and missed parental days at work, were calculated for those with completed parental questionnaires.

Odds ratios for various severity of disease outcomes with respect to RSV colonisation status were estimated in univariate and multivariable logistic regression models and reported using odds ratios (OR), 95% confidence intervals and p-values. Among RSV positive participants odds ratios for factors relating to hospitalisation were estimated in the same way.

### Health economic analysis

#### Healthcare utilisation costs

The total costs associated with managing RSV were derived from the type and number of interactions with the healthcare systems recorded in patient healthcare records, supplemented by any additional interactions noted by parents and caregivers. Costs associated with healthcare resource utilisation were examined, including previous healthcare visits, current healthcare attendance, duration of hospitalisation, level of care, and readmissions. The type of interaction included general practice (GP) consultation (in person or telephone), attendance at GP out-of-hours or walk-in surgeries, ED attendance and hospital admission (general paediatric ward, high-dependency unit, paediatric intensive care unit).

Costs for primary care services were taken from the Health and Social Care 2022 manual(15). Patients attending ED and then discharged home, as well as those who were then admitted (following ED) were costed for the ED attendance. In the latter case, costs were also included for the subsequent admission(s). Costs for ED attendance and hospital admissions (paediatric ward, HDU and PICU) were taken from Hodgson et al(16)

Short-term hospital admission was defined as <3 days; and long stay admissions were defined as > 3 days. In line with published data(16) the cost for a patient admitted > 3 days included both the short and long-stay component, i.e., a single, rather than composite cost was applied. Costs for HDU and PICU admissions were assumed to be the same. For admission cost calculations, total number of days on the ward (taken at day 28), HDU and PICU were included.

Repeat hospital admissions - where the total number of ward days (HDU and PICU) exceeded 3 days - were categorised as long stays. For example, if a patient reported two separate admissions, one lasting two days and the other three days, this healthcare contact was coded as one continuous long stay (incurring the associated cost).

### Work impact

Parents and caregivers reported the number of working hours lost due to RSV (e.g., through taking time off work to care for the child, attend GP consultations etc.) These working hours lost were converted into monetary units (using the Human Capital Approach) by applying the median hourly wage for the UK (£15.88 per hour, Office for National Statistics, 2023(17)) to provide an estimate of the potential economic cost of RSV through lost working hours.

This results in an equivalent, rather than actual loss of earnings.

## 2. Results

### Recruitment summary

A total of 2,074 participants were recruited, with 74 participants withdrawn (Supplementary Fig. 1). After exclusions, the final analysis included 410 participants from primary care (283 primary care, 127 from walk-in centres (WIC)), and 1,590 from secondary or tertiary care. In the URTI sub-study, 220 participants from primary care and 98 from WIC were included in the final analysis (Fig. 1). The overall completion rates for parent/guardian questionnaires were 56.0% at day 14, 53.3% at day 28, and 46.1% for both timepoints (Fig. 1).

### Baseline demographics

Overall, 70.2% of children had no significant underlying co-morbidity or prematurity; 56.9% of participants were male, and 86.9% were of White British ethnicity. 53.7% were <12 months old, 28.8% were 12-<24 months and 17.4% were 24-<36 months (Table 1).

**Table 1.**
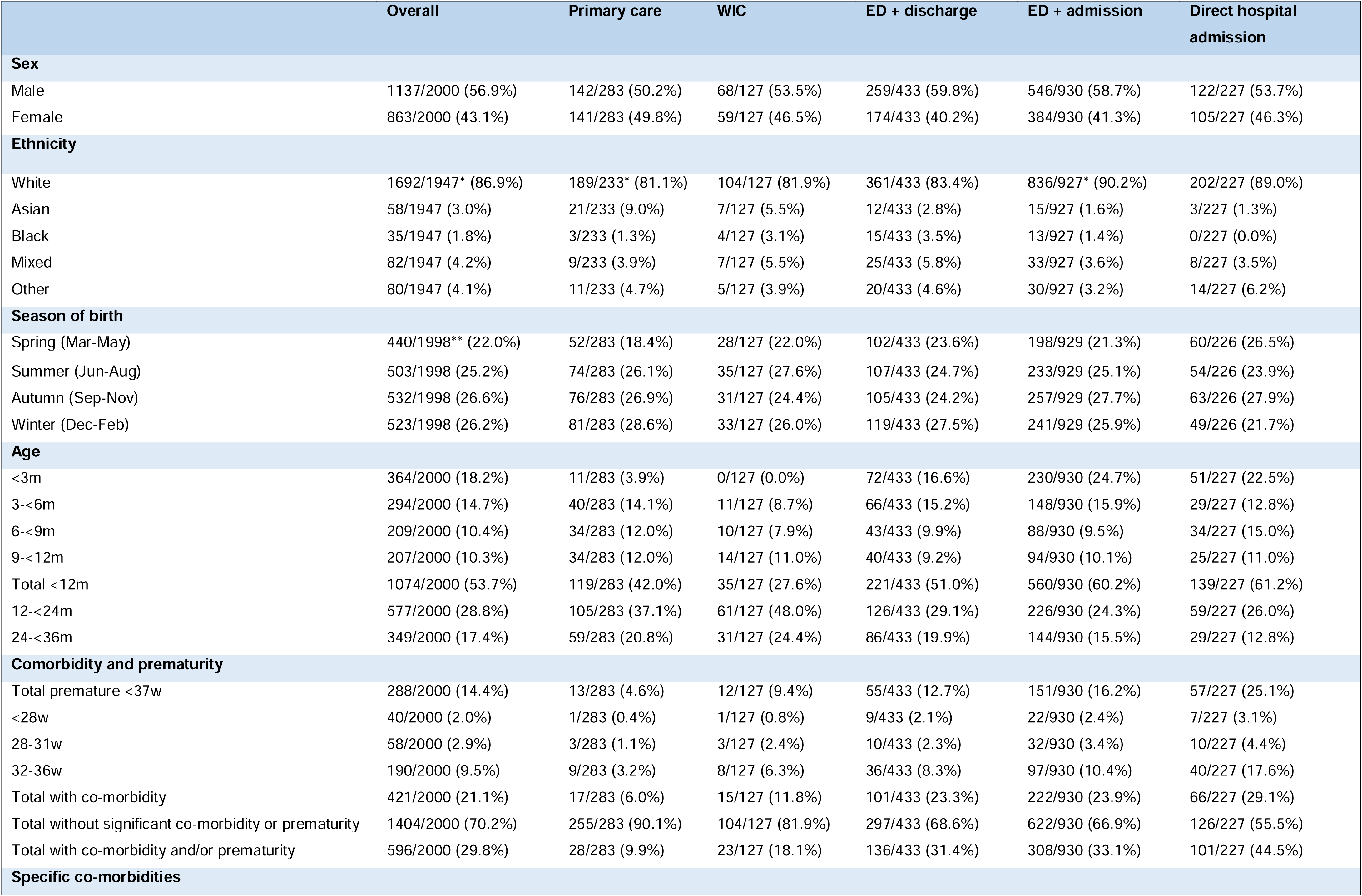
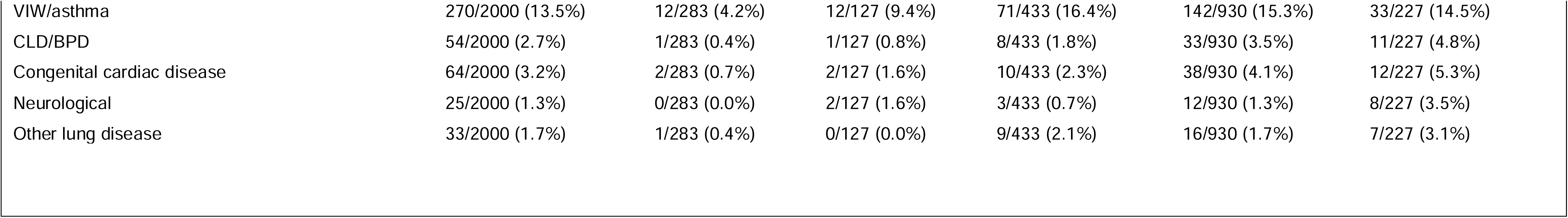
Baseline characteristics of participants according to the healthcare area they were recruited from. ED=emergency department; WIC=walk in centre; PC=primary care; w=weeks; m=months; VIW=viral induced wheeze; CLD=chronic lung disease of prematurity; BPD=bronchopulmonary dysplasia; m = months; w=weeks

|  | Overall | Primary care | WIC | ED + discharge | ED + admission | Direct hospital admission |
| --- | --- | --- | --- | --- | --- | --- |
| **Sex** |  |  |  |  |  |  |
| Male | 1137/2000 (56.9%) | 142/283 (50.2%) | 68/127 (53.5%) | 259/433 (59.8%) | 546/930 (58.7%) | 122/227 (53.7%) |
| Female | 863/2000 (43.1%) | 141/283 (49.8%) | 59/127 (46.5%) | 174/433 (40.2%) | 384/930 (41.3%) | 105/227 (46.3%) |
| **Ethnicity** |  |  |  |  |  |  |
| White | 1692/1947* (86.9%) | 189/233* (81.1%) | 104/127 (81.9%) | 361/433 (83.4%) | 836/927* (90.2%) | 202/227 (89.0%) |
| Asian | 58/1947 (3.0%) | 21/233 (9.0%) | 7/127 (5.5%) | 12/433 (2.8%) | 15/927 (1.6%) | 3/227 (1.3%) |
| Black | 35/1947 (1.8%) | 3/233 (1.3%) | 4/127 (3.1%) | 15/433 (3.5%) | 13/927 (1.4%) | 0/227 (0.0%) |
| Mixed | 82/1947 (4.2%) | 9/233 (3.9%) | 7/127 (5.5%) | 25/433 (5.8%) | 33/927 (3.6%) | 8/227 (3.5%) |
| Other | 80/1947 (4.1%) | 11/233 (4.7%) | 5/127 (3.9%) | 20/433 (4.6%) | 30/927 (3.2%) | 14/227 (6.2%) |
| **Season of birth** |  |  |  |  |  |  |
| Spring (Mar-May) | 440/1998** (22.0%) | 52/283 (18.4%) | 28/127 (22.0%) | 102/433 (23.6%) | 198/929 (21.3%) | 60/226 (26.5%) |
| Summer (Jun-Aug) | 503/1998 (25.2%) | 74/283 (26.1%) | 35/127 (27.6%) | 107/433 (24.7%) | 233/929 (25.1%) | 54/226 (23.9%) |
| Autumn (Sep-Nov) | 532/1998 (26.6%) | 76/283 (26.9%) | 31/127 (24.4%) | 105/433 (24.2%) | 257/929 (27.7%) | 63/226 (27.9%) |
| Winter (Dec-Feb) | 523/1998 (26.2%) | 81/283 (28.6%) | 33/127 (26.0%) | 119/433 (27.5%) | 241/929 (25.9%) | 49/226 (21.7%) |
| **Age** |  |  |  |  |  |  |
| <3m | 364/2000 (18.2%) | 11/283 (3.9%) | 0/127 (0.0%) | 72/433 (16.6%) | 230/930 (24.7%) | 51/227 (22.5%) |
| 3-<6m | 294/2000 (14.7%) | 40/283 (14.1%) | 11/127 (8.7%) | 66/433 (15.2%) | 148/930 (15.9%) | 29/227 (12.8%) |
| 6-<9m | 209/2000 (10.4%) | 34/283 (12.0%) | 10/127 (7.9%) | 43/433 (9.9%) | 88/930 (9.5%) | 34/227 (15.0%) |
| 9-<12m | 207/2000 (10.3%) | 34/283 (12.0%) | 14/127 (11.0%) | 40/433 (9.2%) | 94/930 (10.1%) | 25/227 (11.0%) |
| Total <12m | 1074/2000 (53.7%) | 119/283 (42.0%) | 35/127 (27.6%) | 221/433 (51.0%) | 560/930 (60.2%) | 139/227 (61.2%) |
| 12-<24m | 577/2000 (28.8%) | 105/283 (37.1%) | 61/127 (48.0%) | 126/433 (29.1%) | 226/930 (24.3%) | 59/227 (26.0%) |
| 24-<36m | 349/2000 (17.4%) | 59/283 (20.8%) | 31/127 (24.4%) | 86/433 (19.9%) | 144/930 (15.5%) | 29/227 (12.8%) |
| **Comorbidity and prematurity** |  |  |  |  |  |  |
| Total premature <37w | 288/2000 (14.4%) | 13/283 (4.6%) | 12/127 (9.4%) | 55/433 (12.7%) | 151/930 (16.2%) | 57/227 (25.1%) |
| <28w | 40/2000 (2.0%) | 1/283 (0.4%) | 1/127 (0.8%) | 9/433 (2.1%) | 22/930 (2.4%) | 7/227 (3.1%) |
| 28-31w | 58/2000 (2.9%) | 3/283 (1.1%) | 3/127 (2.4%) | 10/433 (2.3%) | 32/930 (3.4%) | 10/227 (4.4%) |
| 32-36w | 190/2000 (9.5%) | 9/283 (3.2%) | 8/127 (6.3%) | 36/433 (8.3%) | 97/930 (10.4%) | 40/227 (17.6%) |
| Total with co-morbidity | 421/2000 (21.1%) | 17/283 (6.0%) | 15/127 (11.8%) | 101/433 (23.3%) | 222/930 (23.9%) | 66/227 (29.1%) |
| Total without significant co-morbidity or prematurity | 1404/2000 (70.2%) | 255/283 (90.1%) | 104/127 (81.9%) | 297/433 (68.6%) | 622/930 (66.9%) | 126/227 (55.5%) |
| Total with co-morbidity and/or prematurity | 596/2000 (29.8%) | 28/283 (9.9%) | 23/127 (18.1%) | 136/433 (31.4%) | 308/930 (33.1%) | 101/227 (44.5%) |
| **Specific co-morbidities** |  |  |  |  |  |  |
| VIW/asthma | 270/2000 (13.5%) | 12/283 (4.2%) | 12/127 (9.4%) | 71/433 (16.4%) | 142/930 (15.3%) | 33/227 (14.5%) |
| CLD/BPD | 54/2000 (2.7%) | 1/283 (0.4%) | 1/127 (0.8%) | 8/433 (1.8%) | 33/930 (3.5%) | 11/227 (4.8%) |
| Congenital cardiac disease | 64/2000 (3.2%) | 2/283 (0.7%) | 2/127 (1.6%) | 10/433 (2.3%) | 38/930 (4.1%) | 12/227 (5.3%) |
| Neurological | 25/2000 (1.3%) | 0/283 (0.0%) | 2/127 (1.6%) | 3/433 (0.7%) | 12/930 (1.3%) | 8/227 (3.5%) |
| Other lung disease | 33/2000 (1.7%) | 1/283 (0.4%) | 0/127 (0.0%) | 9/433 (2.1%) | 16/930 (1.7%) | 7/227 (3.1%) |

### RSV rates

As expected, RSV positivity varied by month and was in line with UK Health Security Agency (UKHSA) surveillance data for hospitals admissions in the Northwest of England (Fig 2, Supplementary Fig 2, 3), which reports RSV positivity on hospital admissions only (18). RSV positivity varied by care setting, season, age, and presence of LRTI features (Table 2, Supplementary Fig 1, 2).

**Table 2.** RSV positivity by level of care, URTI, LRTI, age, sex, ethnicity, gestation and co-morbidity status. URTI = upper respiratory tract infection; LRTI = lower respiratory tract infection; m=months; WIC=walk in centre; w=weeks;ED=emergency department; m=months; w=weeks

|  | Overall | RSV +ve | RSV -ve |
| --- | --- | --- | --- |
| **Overall** | 2000 (100%) | 796 (39.8%) | 1204 (60.2%) |
| **Level of care** |  |  |  |
| Primary care overall | 283 (14.1%) | 51 (18.0%) | 232 (82.0%) |
| Primary care LRTI | 63 (3.1%) | 23 (36.5%) | 40 (63.5%) |
| Primary care URTI | 220 (11.0%) | 28 (12.7%) | 192 (87.3%) |
| WIC Overall | 127 (6.3%) | 5 (3.9%) | 122 (96.1%) |
| WIC LRTI | 29 (1.5%) | 3 (10.3%) | 26 (89.7%) |
| WIC URTI | 98 (4.9%) | 2 (2.0%) | 96 (98.0%) |
| ED + discharge | 433 (21.6%) | 157 (36.3%) | 276 (63.7%) |
| ED + admission | 930 (46.5%) | 465 (50.0%) | 465 (50.0%) |
| Direct admission | 227 (11.3%) | 118 (52.0%) | 109 (48.0%) |
| Hospital attendances overall | 1,590 (79.4%) | 740 (46.5%) | 850 (53.5%) |
| Hospital admissions overall | 1157 (57.9%) | 583 (50.4%) | 574 (49.6%) |
| HDU admission | 109 (5.5%) | 60 (55.0%) | 49 (45.0%) |
| PICU admission | 66 (3.3%) | 38 (57.6%) | 28 (42.4%) |
| **Sex** |  |  |  |
| Male | 1137 (56.9%) | 450 (39.6%) | 687 (60.4%) |
| Female | 863 (43.1%) | 346 (40.1%) | 517 (59.9%) |
| **Ethnicity** |  |  |  |
| White | 1692 (86.9%) | 699 (41.3%) | 993 (58.7%) |
| Asian | 58 (3.0%) | 16 (27.6%) | 42 (72.4%) |
| Black | 35 (1.8%) | 12 (34.3%) | 23 (65.7%) |
| Mixed | 82 (4.2%) | 33 (40.2%) | 49 (59.8%) |
| Other | 80 (4.1%) | 27 (33.8%) | 53 (66.3%) |
| **Age** |  |  |  |
| <3m | 364 (18.2%) | 214 (58.8%) | 150 (41.2%) |
| 3-<6m | 294 (14.7%) | 162 (55.1%) | 132 (44.9%) |
| 6-<9 | 209 (10.4%) | 80 (38.3%) | 129 (61.7%) |
| 9-<12 | 207 (10.3%) | 71 (34.3%) | 136 (65.7%) |
| Total<12m | 1074 (53.7%) | 527 (49.1%) | 547 (50.9%) |
| 12-<23m | 577 (28.8%) | 172 (29.8%) | 405 (70.2%) |
| 24-<35m | 349 (17.4%) | 97 (27.8%) | 252 (72.2%) |
| **Season of birth** |  |  |  |
| Spring (Mar-May) | 440 (22.0%) | 181 (41.1%) | 259 (58.9%) |
| Summer (Jun-Aug) | 503 (25.2%) | 211 (41.9%) | 292 (58.1%) |
| Autumn (Sep-Nov) | 532 (26.6%) | 238 (44.7%) | 294 (55.3%) |
| Winter (Dec-Feb) | 523 (26.2%) | 165 (31.5%) | 358 (68.5%) |
| **Co-morbidity and prematurity** |  |  |  |
| Significant comorbidity with or without prematurity | 421 (21.1%) | 133 (31.6%) | 288 (68.4%) |
| Prematurity (Gestation <37w) | 288 (14.4%) | 113 (39.2%) | 175 (60.8%) |
| Significant co-morbidity and/or prematurity | 596 (29.8%) | 212 (35.6%) | 384 (64.4%) |
| Significant co-morbidity and/or prematurity excluding palivizumab subgroup | 542 (27.1%) | 202 (37.3%) | 340 (62.7%) |
| Total without significant co-morbidity or prematurity | 1404 (70.2%) | 584 (41.6%) | 820 (58.4%) |

Within hospital settings, RSV positivity was higher in participants admitted via ED or to the hospital directly (50.4%) compared with those who attended ED and were discharged (36.3%) (Chi-squared test p<0.01). In primary care overall RSV positivity was 18.0% with higher positivity in children with LRTI (36.5%) than those without LRTI features (12.7%) (Table 2). WIC RSV positivity was low (3.9%), however WIC recruitment occurred only from January - March 2023 when UKHSA reported RSV rates were low (Fig 1) (18).

### Clinical presentation

RSV-positive participants were significantly more likely to present with features of respiratory distress including shortness of breath (OR 1.32, 95% CI 1.04-1.67, p=0.024), nasal flaring (OR 1.39, 95% CI 1.01-1.89, p=0.040), and apnoea/chest recession/head bobbing (OR 1.25, 95% CI 1.01-1.54, p=0.039) than RSV-negative participants (Supplementary Table 4).

### Repeated healthcare presentations

Multiple presentations to different healthcare settings with the presumed same illness were more common in RSV-positive participants. Prior to recruitment, 59.0% of RSV-positive participants had had face-to-face contact with primary healthcare services compared with 43.8% of RSV-negative participants (Chi-squared test p<0.001, table 3, Supplementary Table 5). Of RSV-positive participants initially discharged 7.9% were subsequently readmitted compared with 6.5% of RSV-negative. (Chi-squared test p=0.218, table 3, Supplementary Table 5).

**Table 3.**
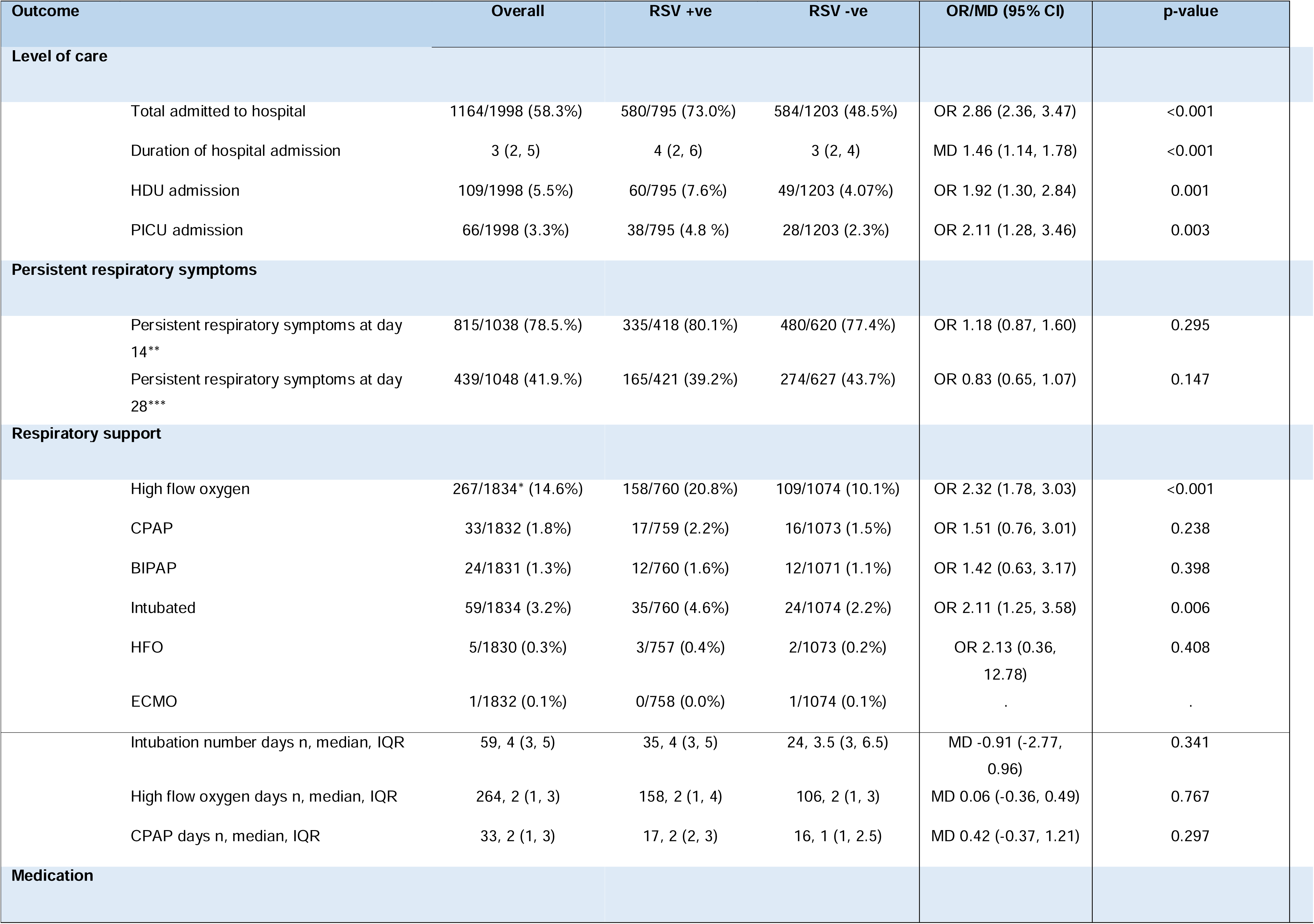
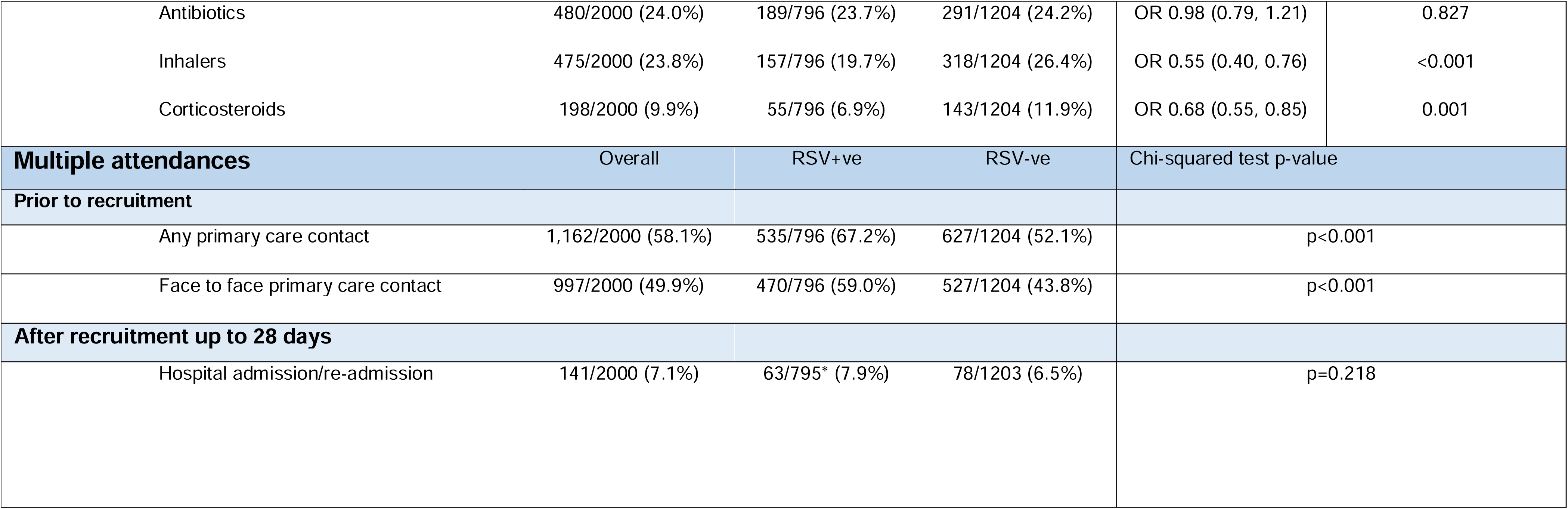
Comparison of severity outcomes, medical interventions and multiple healthcare attendances by RSV status. MD = mean difference, HDU = high dependency unit, PICU = paediatric intensive care unit, VIW = viral induced wheeze, BPD = Bronchopulmonary dysplasia, CLD = chronic lung disease, RR=relative risk, HFL = high flow oxygen, HFO = High frequency oscillation ventilation, ECMO = Extracorporeal membrane oxygenation, CPAP = continuous positive airway pressure, BiPAP = Biphasic positive airway pressure. *missing day 28 follow up data for respiratory support, **denominator is those who had completed day 14 questionnaires, ***denominator is those who had completed day 28 questionnaires

| Outcome |  | Overall | RSV +ve | RSV -ve | OR/MD (95% CI) | p-value |
| --- | --- | --- | --- | --- | --- | --- |
| **Level of care** |  |  |  |  |  |  |
| Total admitted to hospital |  | 1164/1998 (58.3%) | 580/795 (73.0%) | 584/1203 (48.5%) | OR 2.86 (2.36, 3.47) | <0.001 |
| Duration of hospital admission |  | 3 (2, 5) | 4 (2, 6) | 3 (2, 4) | MD 1.46 (1.14, 1.78) | <0.001 |
| HDU admission |  | 109/1998 (5.5%) | 60/795 (7.6%) | 49/1203 (4.07%) | OR 1.92 (1.30, 2.84) | 0.001 |
| PICU admission |  | 66/1998 (3.3%) | 38/795 (4.8 %) | 28/1203 (2.3%) | OR 2.11 (1.28, 3.46) | 0.003 |
| **Persistent respiratory symptoms** |  |  |  |  |  |  |
| Persistent respiratory symptoms at day 14** |  | 815/1038 (78.5%) | 335/418 (80.1%) | 480/620 (77.4%) | OR 1.18 (0.87, 1.60) | 0.295 |
| Persistent respiratory symptoms at day 28*** |  | 439/1048 (41.9%) | 165/421 (39.2%) | 274/627 (43.7%) | OR 0.83 (0.65, 1.07) | 0.147 |
| **Respiratory support** |  |  |  |  |  |  |
| High flow oxygen |  | 267/1834* (14.6%) | 158/760 (20.8%) | 109/1074 (10.1%) | OR 2.32 (1.78, 3.03) | <0.001 |
| CPAP |  | 33/1832 (1.8%) | 17/759 (2.2%) | 16/1073 (1.5%) | OR 1.51 (0.76, 3.01) | 0.238 |
| BIPAP |  | 24/1831 (1.3%) | 12/760 (1.6%) | 12/1071 (1.1%) | OR 1.42 (0.63, 3.17) | 0.398 |
| Intubated |  | 59/1834 (3.2%) | 35/760 (4.6%) | 24/1074 (2.2%) | OR 2.11 (1.25, 3.58) | 0.006 |
| HFO |  | 5/1830 (0.3%) | 3/757 (0.4%) | 2/1073 (0.2%) | OR 2.13 (0.36, 12.78) | 0.408 |
| ECMO |  | 1/1832 (0.1%) | 0/758 (0.0%) | 1/1074 (0.1%) | . | . |
| Intubation number days n, median, IQR |  | 59, 4 (3, 5) | 35, 4 (3, 5) | 24, 3.5 (3, 6.5) | MD -0.91 (-2.77, 0.96) | 0.341 |
| High flow oxygen days n, median, IQR |  | 264, 2 (1, 3) | 158, 2 (1, 4) | 106, 2 (1, 3) | MD 0.06 (-0.36, 0.49) | 0.767 |
| CPAP days n, median, IQR |  | 33, 2 (1, 3) | 17, 2 (2, 3) | 16, 1 (1, 2.5) | MD 0.42 (-0.37, 1.21) | 0.297 |
| **Medication** |  |  |  |  |  |  |

**Table 3 Comparison of severity outcomes, medical interventions and multiple healthcare attendances by RSV status** MD = mean difference, HDU = high dependency unit, PICU = paediatric intensive care unit, VIW = viral induced wheeze, BPD = Bronchopulmonary dysplasia, CLD = chronic lung disease, RR=relative risk, HFL = high flow oxygen, HFO = High frequency oscillation ventilation, ECMO = Extracorporeal membrane oxygenation, CPAP = continuous positive airway pressure, BiPAP = Biphasic positive airway pressure.
|  |  |  |  |  |  |
| --- | --- | --- | --- | --- | --- |
| Antibiotics | 480/2000 (24.0%) | 189/796 (23.7%) | 291/1204 (24.2%) | OR 0.98 (0.79, 1.21) | 0.827 |
| Inhalers | 475/2000 (23.8%) | 157/796 (19.7%) | 318/1204 (26.4%) | OR 0.55 (0.40, 0.76) | <0.001 |
| Corticosteroids | 198/2000 (9.9%) | 55/796 (6.9%) | 143/1204 (11.9%) | OR 0.68 (0.55, 0.85) | 0.001 |
| **Multiple attendances** | **Overall** | **RSV+ve** | **RSV-ve** | **Chi-squared test p-value** |  |
| **Prior to recruitment** |  |  |  |  |  |
| Any primary care contact | 1,162/2000 (58.1%) | 535/796 (67.2%) | 627/1204 (52.1%) | p<0.001 |  |
| Face to face primary care contact | 997/2000 (49.9%) | 470/796 (59.0%) | 527/1204 (43.8%) | p<0.001 |  |
| **After recruitment up to 28 days** |  |  |  |  |  |
| Hospital admission/re-admission | 141/2000 (7.1%) | 63/795* (7.9%) | 78/1203 (6.5%) | p=0.218 |  |

### Comparison of outcomes by RSV positivity

For most measures of severity, outcomes were worse in RSV-positive participants compared with RSV-negative (Fig 2, Table 3). 73.0% of RSV-positive participants were hospitalised compared with 48.5% of RSV-negative participants (OR 2.86,95% CI 2.36-3.47, <0.001), and median duration of hospital admission was longer (4 days v 3 days, p<0.001). A higher proportion of RSV-positive participants were admitted to HDU (7.6% v 4.1%, OR 1.92, 95% CI 1.30-2.84, p=0.001) or PICU (4.8% v 2.3% , OR 2.11, 95% CI 1.28 - 3.46, p=0.003). Persistence of respiratory symptoms in RSV-positive participants was 80.1% at day 14 and 39.2% at day 28, but there was no statistically significant difference compared with RSV-negative participants: day 14 (OR 1.18, 95% CI 0.87-1.60, p=0.295); day 28 (OR 0.83, 95% CI 0.65-1.07, p=0.147). A high proportion of RSV-positive and RSV-negative participants were prescribed antibiotics (23.7% v 24.2%, OR 0.98, 95% CI 0.79 - 1.21, p=0.827).

### Risk factors for hospitalisation, HDU/PICU admission and medical interventions in RSV-positive participants

Of all hospitalised RSV-positive participants, the majority had no significant underlying co-morbidity or prematurity (70.1%) (Fig 3). Furthermore, of the children admitted to HDU and PICU, 55.0% and 42.1% respectively had no significant underlying co-morbidity or prematurity. Nevertheless, the odds of hospitalisation were increased in certain groups (Fig 4, Supplementary Table 6).

On multivariate analysis, there were increased odds of hospitalisation in children of all levels of prematurity compared to hospitalisation in children born at full term (86.7% v 70.7%, OR 2.24, 95% CI 1.24 - 4.03, p=0.007), including moderate to late prematurity compared to children not born premature on single variable analysis (71.6% v 83.9%, OR 2.16, 95% CI 1.19 - 3.92, p=0.011). Furthermore, there were increased odds of hospitalisation in children <3 months old compared to children in the oldest age category (90.1% v 66.7%, OR 6.61 (3.45, 12.68), p<0.001) and children 3-<6months compared to the oldest age group (OR 1.98, CI 1.10 - 3.55, p=0.022) (Fig 4, Supplementary Table 6).

On multivariate analysis, the odds of admission to HDU/PICU was increased in children born premature (OR 3.86, 95% CI 2.22-6.72, p<0.001) compared to children born at full term including moderate – late premature (OR 3.95, 95% CI 2.30 - 6.77, p<0.001, single variable analysis), children <3months old compared to the oldest age group (OR 19.61 (4.60, 83.51), p<0.001), and children with congenital cardiac disease compared to those without (OR 4.37, 95% CI 1.58-12.10, p=0.004) (Supplementary Fig 5, Supplementary table 8).

### RSV viral co-infection

Co-infection with two or more respiratory viruses was more common in RSV-positive participants (40.8%) than RSV-negative participants (18.3%) and was more common in older age groups (Fig 5, Supplementary Table 12). The most common virus involved in RSV co-infection was Rhinovirus followed by Adenovirus (Fig 5).

### Healthcare economic analysis

Approximately 15% of patients (n=297; 251 RSV-negative; 46 RSV-positive) reported no costing data. Data were available for 1696 participants (85% of the total; 949 RSV-negative [56%]; 747 RSV-positive [44%]). There was a higher healthcare cost in RSV-positive participants (mean per participant = £1811, 95%CI = £1720 to £1903) compared to RSV-negative (mean per participant = £1295, 95% CI = £1220 to £1370), with a total cost of £1,353,144 in RSV-positive and £1,228,554 in RSV-negative. Therefore, there was an average additional treatment cost of £516 per participant and a total additional cost of £124,590 associated with treating RSV-positive participants (Supplementary File 2, Table 2).

### Work Impact

Overall, 601 parents and caregivers reported lost work hours due to their child’s ill health. On average, respondents indicated they lost 32 working hours or approximately four working days. Those with children who were RSV-positive (mean = 32.5 hours; SD = 26.62 hours) and RSV-negative (mean = 32.27 hours; SD = 33.62 hours) displayed a similar loss of working hours. The approximate economic cost to parents was equivalent to an average of £515.47 in RSV-positive and £512.51 in RSV-negative. The total number of lost working hours was 19,445, an equivalent of £308,779. (Supplementary file 2).

## 3. Discussion

Our study demonstrates a substantial RSV burden across all levels of healthcare and provides important data on the burden in primary care and EDs. RSV positivity varied by level of care, with highest positivity in children admitted to hospital (50.4%), and similar RSV positivity in children discharged from ED (36.3%) and primary care LRTI (36.5%). Multiple presentations to different healthcare providers with the same RSV illness were common with 59.0% of participants presenting to the hospital having already consulted face-to-face with primary care. Furthermore, we found a high RSV burden in healthcare settings from previously healthy term-born children, who accounted for 73.3% of medically attended RSV infection, 70.1% of hospitalised children, and 42.1% of children admitted to PICU with RSV. Importantly, we found that even children born moderately preterm were at increased risk of severe disease. These children may not benefit from full protection from maternal vaccination, and many would not qualify for Nirsevimab under current UK guidance. Finally, we demonstrate high healthcare costs associated with RSV as well as the wider economic impact on caregiver working hours lost.

Our study demonstrates a higher prevalence (39.8%) of RSV infection in children <3 years old than described in previous literature (1, 11)(Fig 2). This difference is likely due to study population differences: whilst the European study (RESCEU) included healthy term-born infants only, we included those with co-morbidities and prematurity to represent the whole population who may benefit from preventive therapies and to investigate risk factors for severe disease. Furthermore, our study predominantly recruited those with LRTI features only, unlike previous studies (1,6).

We present important data on the RSV burden in primary care and ED discharges as it is not routine practice to conduct viral nasal swabs in these settings in the UK. Our results are comparable with previous estimates of 20% to 50% of respiratory infections attributable to RSV in primary care (19) (5). Overall RSV rates were likely lower in primary care, as we also included participants with upper respiratory tract infection. Interestingly, the positivity of RSV LRTI in primary care (36.5%) was similar to the positivity in ED discharges (36.3%) which suggests that some children who attend ED with LRTI symptoms, may be suitable for primary care management reflecting pressures on primary care. Further qualitative data is needed to explore this potentially avoidable ED burden. Overall, these data support the use of preventative vaccination to reduce not only the healthcare burden associated with RSV hospitalisations, but also the burden on overstretched primary care and ED services.

We have demonstrated the highest burden of medically attended RSV infection was in healthy term born children, adding to the growing body of evidence that a large proportion of patients with RSV related admissions have no pre-existing risk factors(1, 11). In keeping with other studies(12, 13, 20), we identified that age < 3months, any level of prematurity, congenital cardiac disease and chronic lung disease increased the odds of severe RSV outcomes.

Our study provides insight into the wide-reaching economic impact of RSV infection. We collected data on healthcare attendances throughout the course of illness and found multiple attendances as well as the high cost of hospital admissions contributes to the healthcare economic burden) (Supplementary File 2, Table 1 and 2). This figure is likely an underestimation, as it does not consider subsequent ED or Primary care attendances (only hospital admissions). Furthermore, although there was no difference between RSV-positive and negative participants, the wider economic impact of childhood LRTI is demonstrated by parents/caregivers losing an average of 4 working days per childhood illness.

We have demonstrated that a high proportion of RSV-positive participants were prescribed antibiotics (23.7%). We hypothesise that preventing RSV disease in young children could reduce antibiotic use and contribute to fighting the global threat of antimicrobial resistance.

Finally, this study shows an overall frequency of viral co-infection of 40.8% in RSV-positive participants which is higher than in previous studies (Supplementary Table 10) (1,6).

However, this finding could be explained by the different study populations as mentioned previously.

One limitation of our study - that may introduce bias when comparing RSV-positive and negative outcomes - is that recruitment was permitted when swab results were already known. Another limitation is that our study adopted a pragmatic approach, recruiting from healthcare settings once children present with RTI features. Accordingly, RSV prevalence estimates based on this population will be less accurate than examining a birth cohort study. Finally, there is a risk of reporting bias of parental working days lost as parental questionnaire completion rates were 56.0% at day 14 and 53.3% day 28. Nevertheless, our recruitment model facilitated an analysis of a large cohort of RSV-positive participants, allowing a detailed exploration of the burden on health services, risk factors for severe disease and clinical course over a 28-day period.

Our data suggest that there is a substantial RSV burden in outpatient and inpatient settings and that routine vaccination programmes for RSV will be beneficial in a wide population, including children with no pre-existing risk factors. We have identified increased risk of severe disease in children of any level of prematurity, who may not benefit from full protection from maternal vaccination, we therefore recommend broadening current UK eligibility for Nirsevimab to include this population. In summary, this study provides evidence to support the universal immunisation programme to protect infants against RSV in the UK, but also to widen current eligibility for immunisation of infants with monoclonal antibody against RSV.

## Summary of Conflict of Interest as declared by authors on ICMJE disclosure forms

Funding for this study was provided to the Liverpool School of Tropical Medicine (LSTM) by a Sanofi investigator led grant.

Dr Emma Carter is an employee of LSTM and provided support to the study as funded from the Sanofi grant to LSTM. She also received personal funding from Sanofi to attend the European Society of Clinical Microbiology and Infectious Diseases Global conference.

Dr Andrea Collins is an employee of LSTM and provided support to the study as funded from the Sanofi grant to LSTM. She also received consultancy fees from Sanofi RSV and Sanofi flu. She received funding from Sanofi to attend the European Society conference in Paediatric Infectious Diseases 2023. Unrelated to this study, she has received payment/honoraria for Pfizer European Respiratory Society lecture, sponsorship for North West Thoracic society also arranged from various companies. She has received grants from Pfizer, MSD, CEPI unrelated to this study. She is a member of the Scientific leadership group, The Pandemic Institute.

Mathieu Bangert, Rolf Kramer and Natalya Vassilouthis are employees of Sanofi and have stock/stock options in Sanofi.

Paul McNamara has received consultancy fees from Sanofi for work regarding RSV immunoprophylaxis (personal and institutional).

Helen Hill, Carla Solórzano, Lauren Kerruish, Lauren McLellan, James Dodd, Gregory S.J. Duncan, Kelly Davies, Paula Saunderson, Raqib Huq and Daniela Ferreira are employees of LSTM and provided support to the study as funded by Sanofi but did not receive direct, personal or professional benefits beyond the terms of employment. They have no other conflicts of interest to declare.

Funding to primary care and hospital sites was provided from the Sanofi grant by LSTM for implementation of the study. Shrouk Messahel, Caroline Burchett, Stephen Brearey, Jolanta Bernatoniene, Sudeshna Bhowmik, James Perry, Rebecca Sinfield, Pat Mottram, Paul McNamara, David Lewis, Nadja Van Ginneken and Paul Lipton are employees of these primary and hospital organisations but did not receive any direct, professional or personal benefits beyond the terms of employment. They have no other Conflicts of Interest to declare.

YHEC was commissioned by LSTM to review and analyze the clinical and health economic data presented in the manuscript and staff affiliated with YHEC offered feedback throughout the manuscript development phase. Damian Lewis, Adam B Smith and Andria Collins are employees of YHEC, they did not receive any direct financial, professional or personal benefits beyond the terms of their employment.

Patricia Gonzalez-Dias and Simon B Drysdale are employees of the University of Oxford and provided support in manuscript preparation but did not receive any funding for this and have no other Conflicts of Interest to declare.

Maia Lesosky has an Academy of Medical Sciences Professorship receiving personal funding support for the fellowship which is unrelated to this study.

Shrouk Messahel is a co-applicant for an NIHR Award (162027) A randomised trial of aminophylline, magnesium sulfate or salbutamol intravenous therapy for acute severe asthma in children and young people and has an unpaid voluntary role for PERUKI Vice Chair (Paediatric Emergency Research in UK and Ireland research network), these are unrelated to this study.

## Data Availability

All data produced in the present study are available upon reasonable request to the authors

---

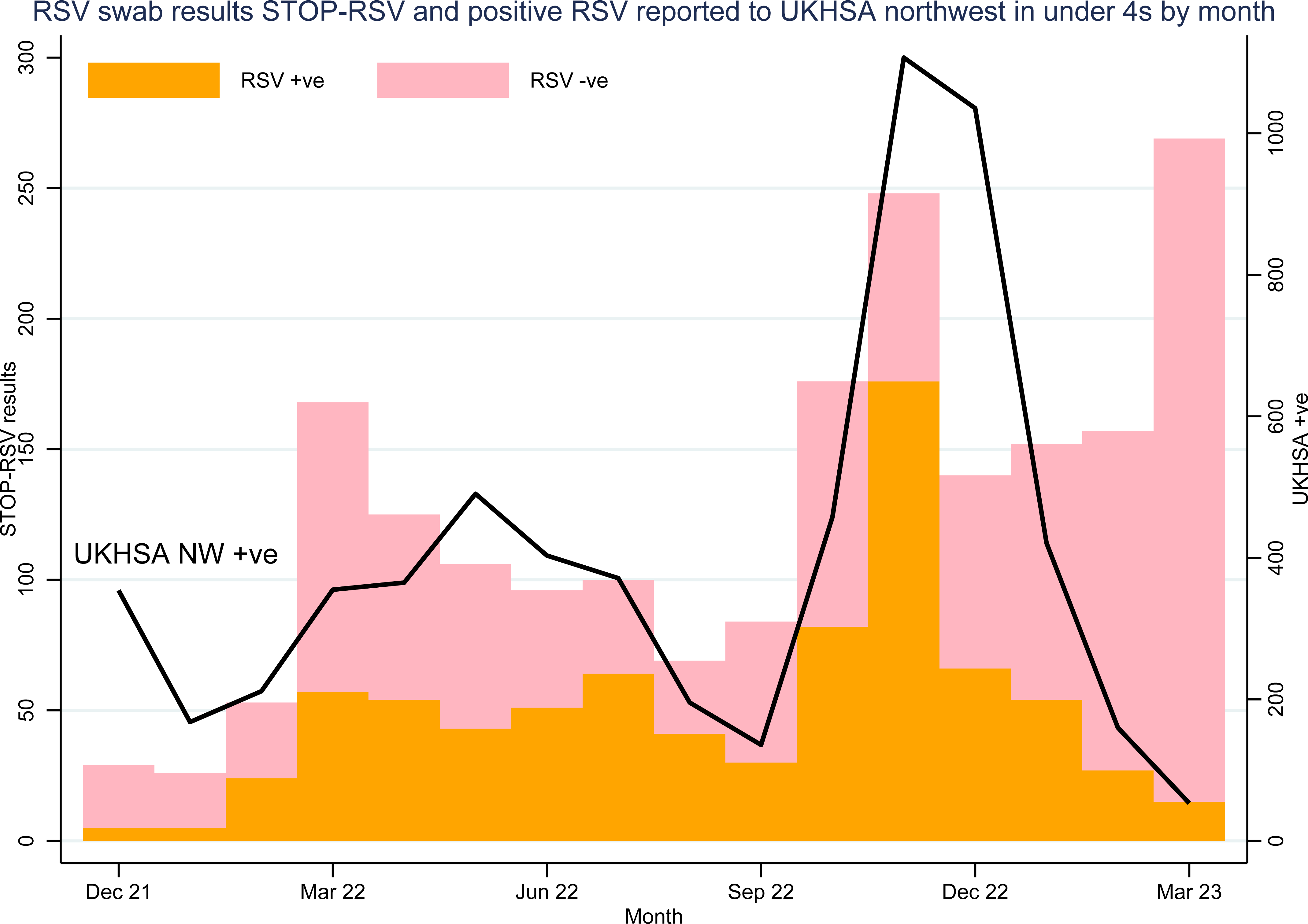

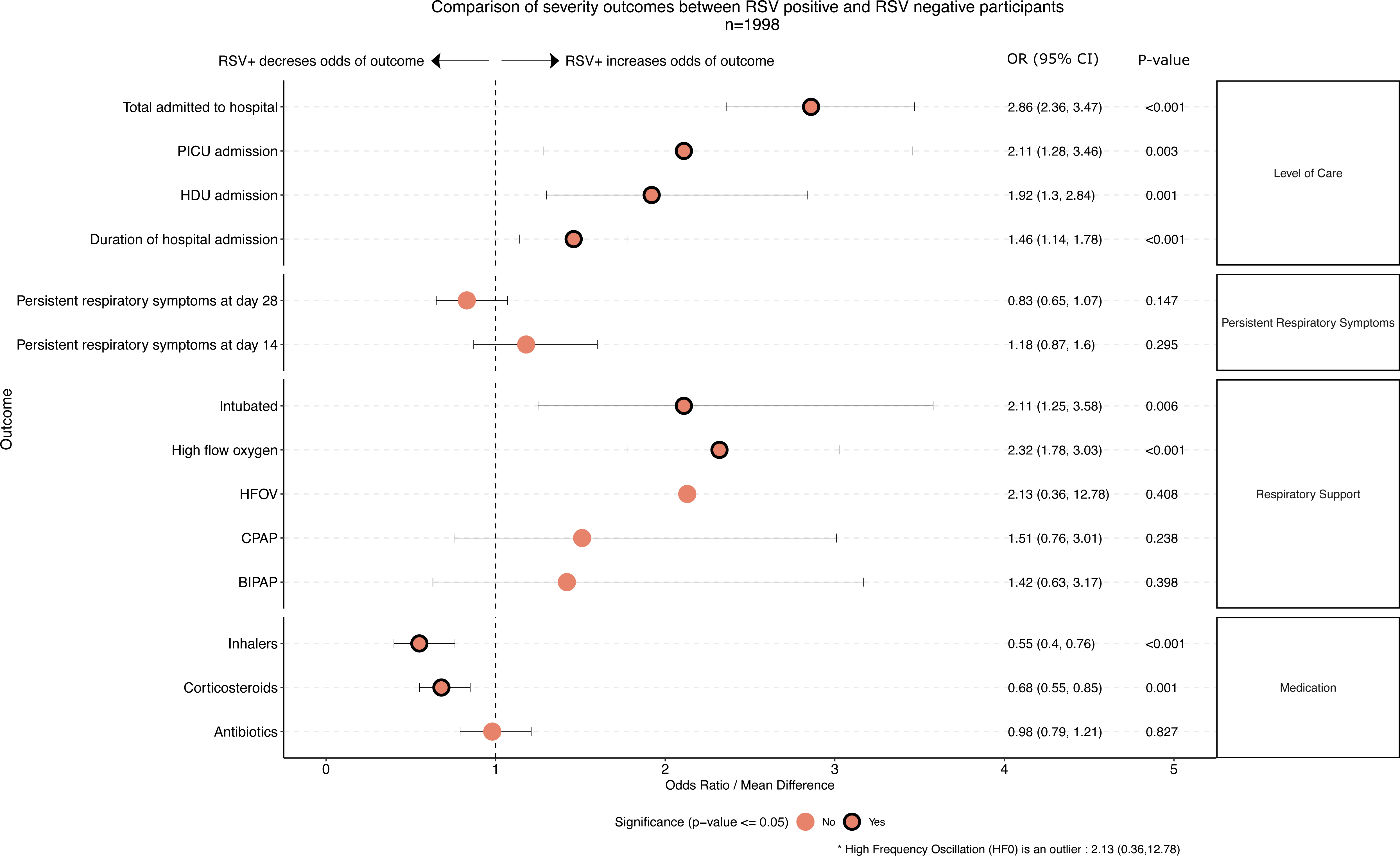

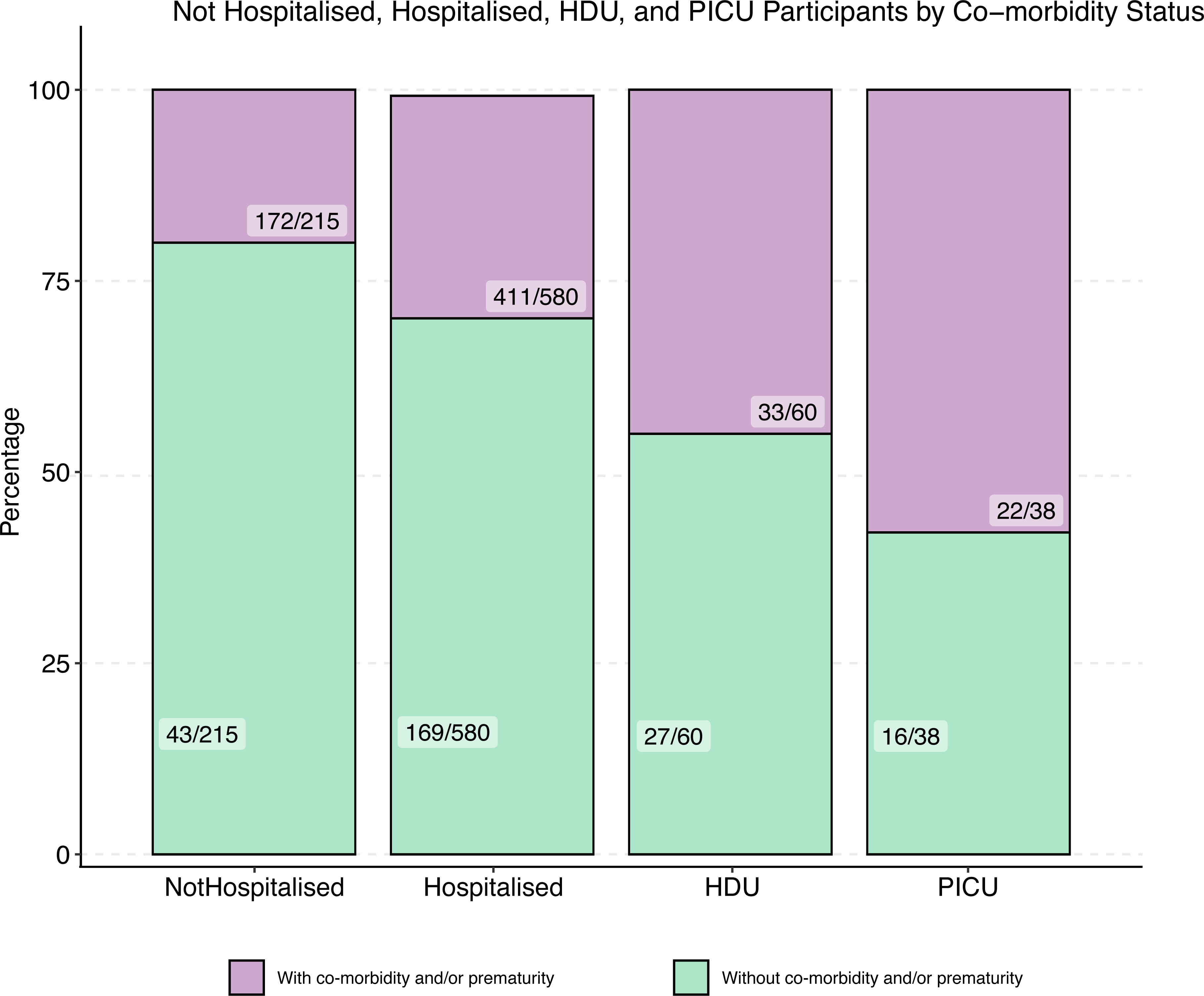

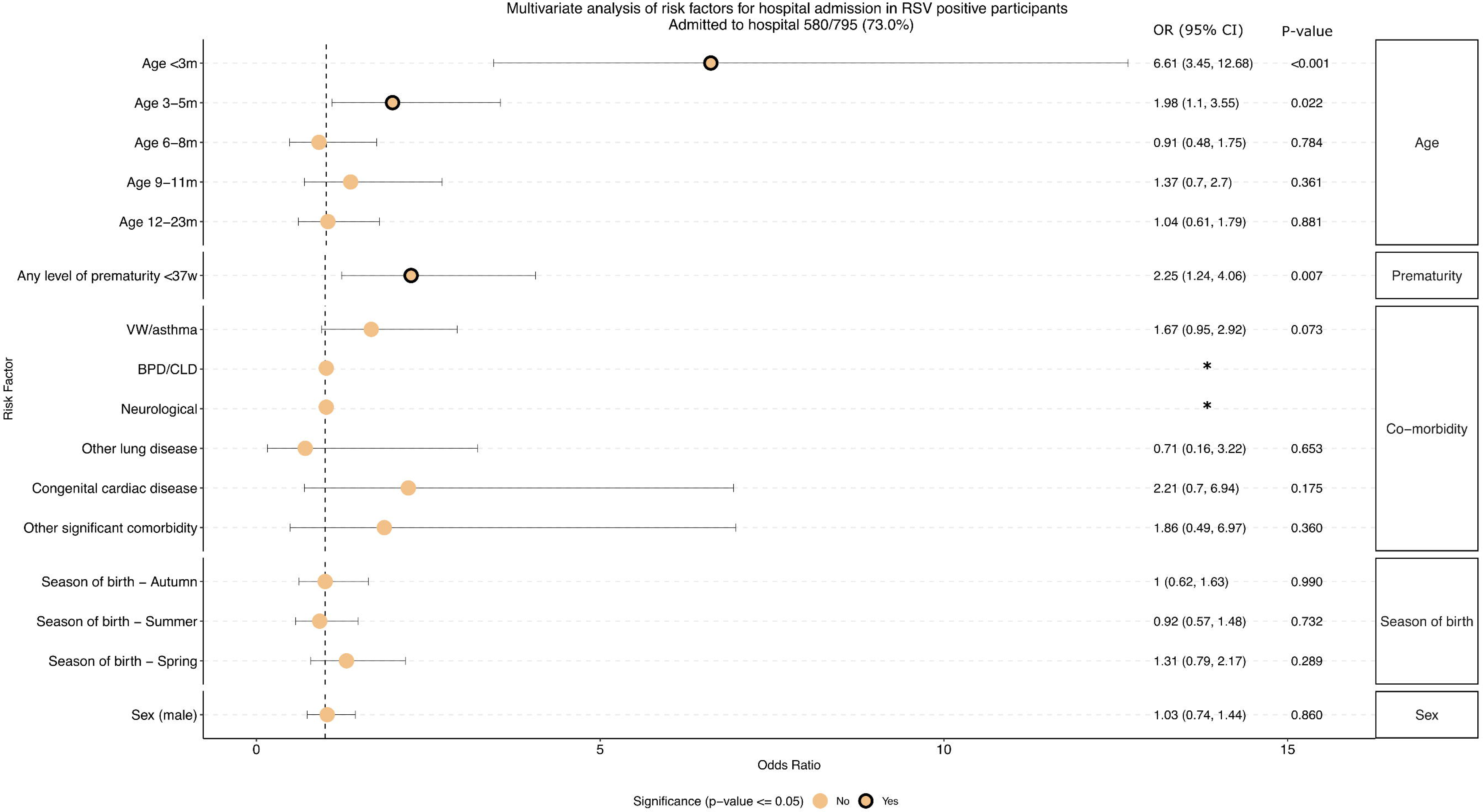

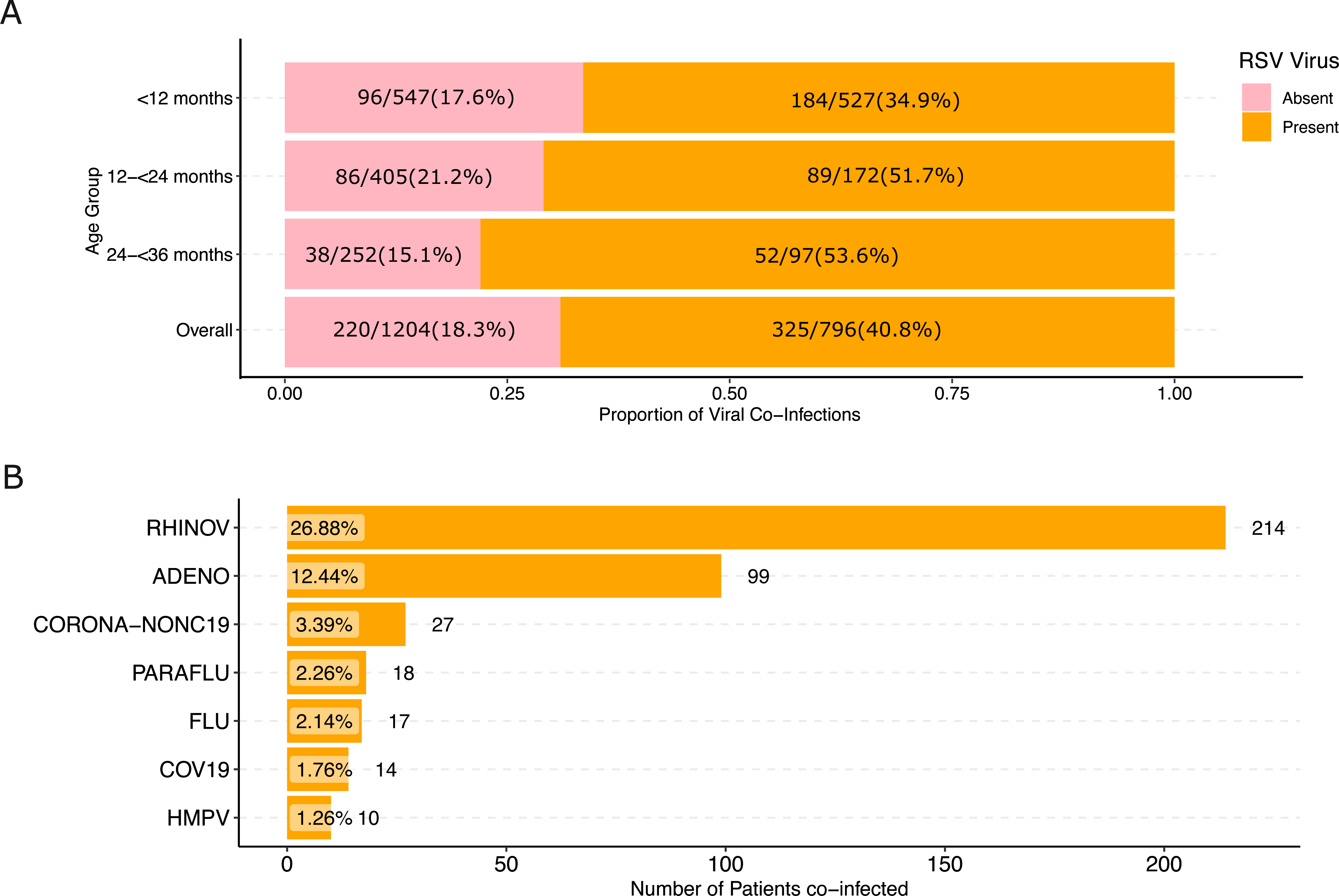

