## Supplementary for "High Respiratory Syncytial Virus Burden in Children Under 3 Years of Age Across All Care Levels in England"

| **Respiratory tract infection symptom**  **(at least one)** | | **Feature of lower respiratory tract infection**  **(at least one)** | |
| --- | --- | --- | --- |
| - Feels hot *or* temperature of >37.8°C - Cough - Nose: Snotty, stuffy, blocked - Earache - Sore throat - Sneezing | AND | | - Audible wheezing (without auscultation) - Shortness of breath, rapid, or shallow breathing for age - Oxygen saturations <94% on air - Crackles, wheeze, or diminished breath sounds on auscultation - Respiratory distress: apnoea, nasal flaring, chest recession, grunting, head bobbing - Central cyanosis - History of above symptoms if intubated prior to site admission |

**Supplementary table 1:** Eligibility criteria for STOP RSV study

| **Panther Fusion Panels**:  Qualitative detection and differentiation**:**  **The Panther Fusion SARS-CoV-2/Flu A/B/RSV assay**  SARS-CoV-2  influenza A and B virus  Respiratory syncytial virus  **The Panther Fusion AdV/hMPV/RV assay**  Adenovirus  Human metapneumovirus  Rhinovirus  **The Panther Fusion Paraflu assay**  Parainfluenza 1, 2, 3, 4 virus | **BIOFIRE System Panel:**  •Adenovirus  •Coronavirus (Non-SARS) 229E, HKU1, NL63, OC43  •SARS-CoV-2  •Human Metapneumovirus  •Human Rhinovirus/Enterovirus  •Influenza A virus  •Influenza B virus  •Parainfluenza virus 1, 2, 3, 4  •Respiratory syncytial virus |
| --- | --- |

**Supplementary table 2**: Panels used for respiratory virus detection from nasal swabs

37/51 (72.5%) Day 14 questionnaire

40/51 (78.4%) Day 28 questionnaire

36 (70.6%) Both

5/5 (100%) Day 14 questionnaire

5/5 (100%) Day 28 questionnaire

89/157 (56.7%) Day 14 questionnaire

82/157 (52.2%) Day 28 questionnaire

72/157 (45.8%) Both

260/465 (55.9%) Day 14 questionnaire

235/465 (50.5%) Day 28 questionnaire

212/465 (45.6%) Both

74/118 (62.7%) Day 14 questionnaire

74/118 (62.7%) Day 28 questionnaire

66/118 (55.9%) Both

51/283 (18%) RSV positive

28/51 (12.7%) URTI

23/51 (36.5%) LRTI

5/127 (3.9%) RSV positive

2/5 (2%) URTI

3/5 (10.3%) LRTI

157/433 (36.3%) RSV positive

465/930 (50.0%) RSV positive

118/227 (52.0%) RSV positive

127 Walk-in-centre

98 RTI without LRTI 29 LRTI

283 General Practice

220 RTI without LRTI

63 LRTI

433 ED discharge

433 LRTI

930 ED admission

930 LRTI

227 Direct hospital admission

227 LRTI

2074 participants recruited

74 withdrawn

- 49 RSV not tested/swab not performed
- 24 Consent withdrawn/invalid
- 1 Age 3 years or over

Primary Care (Total 410)

Hospital (Secondary or Tertiary) (Total 1590)

**Supplementary fig 1** Summary of recruitment, RSV positivity and completion of study activities

|  | **Total Age<12m** | **Age 12-<24m** | **Age 24-<36m** |
| --- | --- | --- | --- |
| **Significant co-morbidity** | 54/527 (10.2%) | 39/172 (22.7%) | 40/97 (41.2%) |
| **Prematurity (<37 weeks)** | 76/527 (14.4%) | 22/172 (12.8%) | 15/97 (15.5%) |
| **Significant co-morbidity or prematurity** | 113/527 (21.4%) | 54/172 (31.4%) | 45/97 (46.4%) |

**Supplementary table 3** RSV positive participants co-morbidity and prematurity status by age

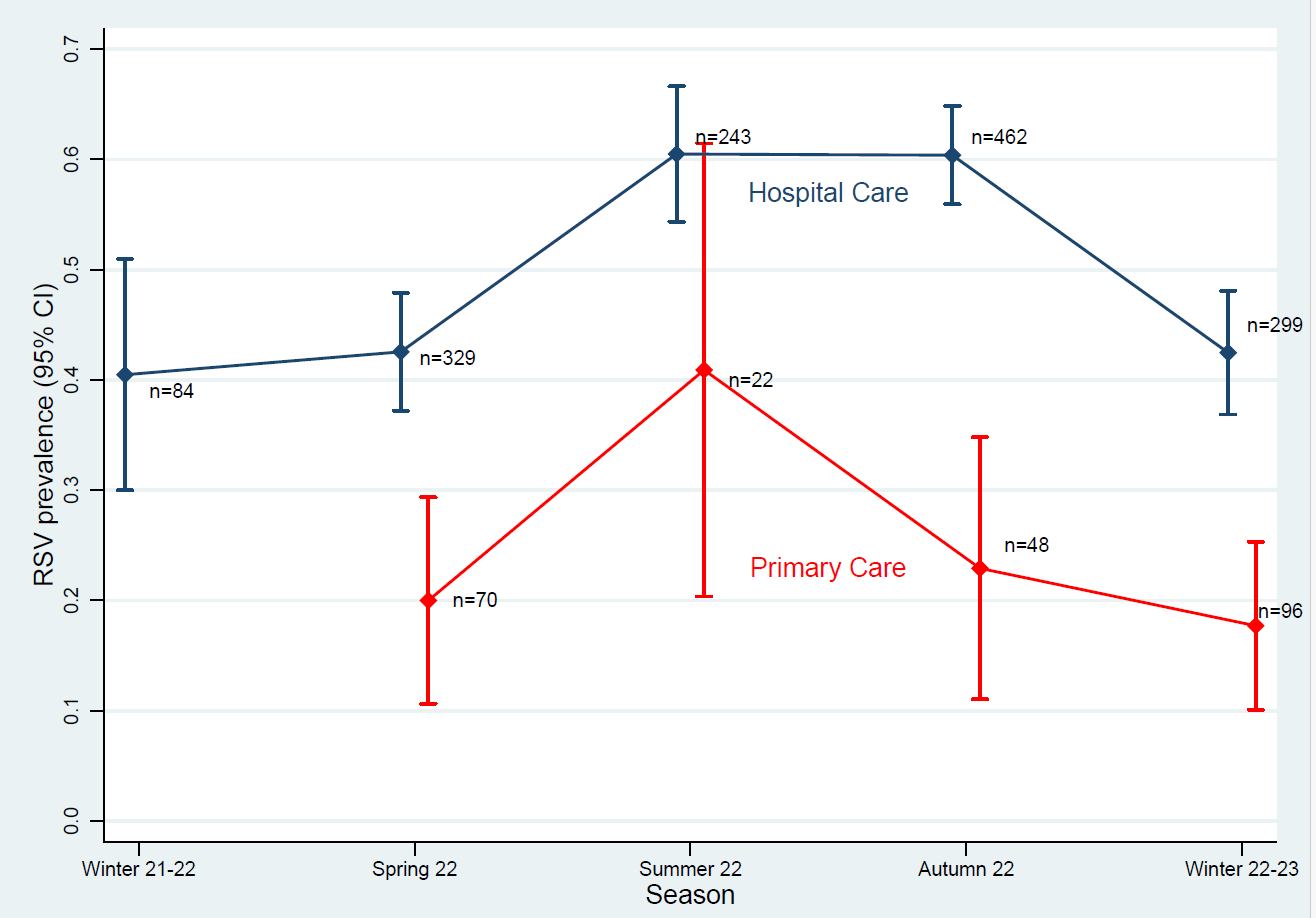

Primary care sites recruitment commenced

Hospital recruitment commenced

**Supplementary Fig 2** RSV positivity rates with confidence intervals across seasons by level of care. Note recruiting sites opened at different times during the study period.

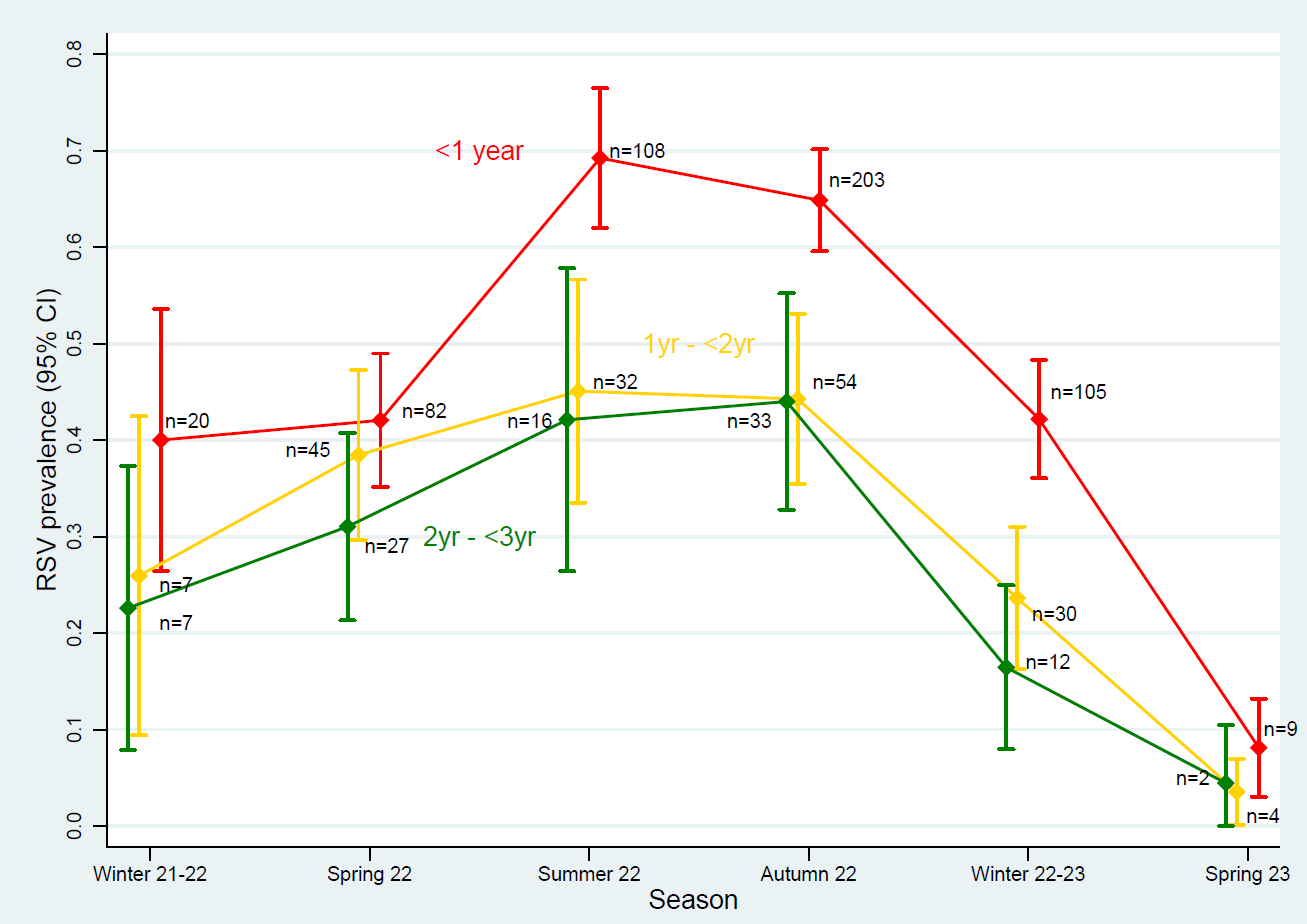

**Supplementary Fig 3 RSV positivity rates with confidence intervals across seasons by age group**

|  |  | **Overall** | **All levels of care RSV +ve** | **All levels of care RSV -ve** | **OR (95% CI)** | **p-value** |
| --- | --- | --- | --- | --- | --- | --- |
| **Non-severe symptoms** | |  |  |  |  |  |
|  | Fever | 956/2000 (47.8%) | 382/796 (48.0%) | 574/1204 (47.7%) | 1.01 (0.85-1.21) | p=0.890 |
|  | Cough | 1775/2000 (88.8%) | 736/796 (92.5%) | 1039/1204 (86.3%) | 1.95 (1.43-2.66) | P<0.001 |
|  | Stuffy/blocked nose | 1619/2000 (81.0%) | 665/796 (83.5%) | 954/1204 (79.2%) | 1.33 (1.05-1.68) | p=0.017 |
|  | Earache | 257/2000 (12.9%) | 75/796 (9.4%) | 182/1204 (15.1%) | 0.58 (0.44-0.78) | p=0.000 |
|  | Sore throat | 511/2000 (25.6%) | 201/796 (25.3%) | 310/1204 (25.7%) | 0.97 (0.79-1.20) | p=0.803 |
|  | Sneezing | 615/2000 (30.8%) | 240/796 (30.2%) | 375/1204 (31.1%) | 0.95 (0.79-1.16) | p=0.637 |
| **Severe symptoms** | |  |  |  |  |  |
|  | SOB/rapid RR | 1329/1682 (79.0%) | 624/766 (81.5%) | 705/916 (77.0%) | 1.32 (1.04-1.67) | p=0.024 |
|  | Audible wheeze without auscultation | 518/1682 (30.8%) | 224/766 (29.2%) | 294/916 (32.1%) | 0.87 (0.71-1.08) | p=0.207 |
|  | Sats<94% on air | 404/1682 (24.0%) | 188/766 (24.5%) | 216/916 (23.6%) | 1.13 (0.90-1.43) | p=0.291 |
|  | Crackes, wheeze or diminished breath | 1003/1682 (59.6%) | 474/766 (61.9%) | 529/916 (57.8%) | 1.19 (0.98-1.44) | p=0.086 |
|  | Respiratory distress | 1174/1682 (69.8%) | 554/766 (72.3%) | 620/916 (67.7%) | 1.25 (1.01-1.54) | p=0.039 |
|  | Nasal flaring | 178/1682 (10.6%) | 94/766 (12.3%) | 84/916 (9.2%) | 1.39 (1.01-1.89) | p=0.040 |
|  | Central cyanosis | 61/1682 (3.6%) | 27/766 (3.5%) | 34/916 (3.7%) | 0.95 (0.57-1.59) | p=0.838 |

**Supplementary table 4 Baseline clinical presentation by RSV status** URTI = upper respiratory tract infection; LRTI = lower respiratory tract infection; SOB = shortness of breath; RR=respiratory rate. *Hospital includes emergency department + discharge, emergency department + admission and direct hospital admission

|  |  |  |  | | | |
| --- | --- | --- | --- | --- | --- | --- |
|  | **Overall RSV+ (number of participants)** | **Overall RSV-** | **Primary care/WIC** | **Hospital sites overall** | **ED+discharge** | **Hospital admission (direct or via ED)** |
| Prior to recruitment | | | | | | |
| Any PC contact, n/N (%) | 535/796 (67.2%) | 627/1204 (52.1%) | 7/56 (12.5%) | 528/740 (71.4%) | 112/157 (71.3%) | 416/583 (71.4%) |
| Any face to face PC contact | 470/796 (59.0%) | 527/1204 (43.8%) | 4/56 (7.1%) | 466/740 (63.0%) | 97/157 (61.8%) | 369/583 (63.3%) |
| GP face to face consultation, n/N (%) | 291/796 (36.6%) | 279/1204 (23.2%) | 2/56 (3.6%) | 289/740 (39.1%) | 65/157 (41.4%) | 224/583 (38.4%) |
| GP out of hours, n/N (%) | 153/796 (19.2%) | 218/1204 (18.1%) | 1/56 (1.8%) | 152/740 (20.5%) | 22/157 (14.0%) | 130/583 (22.3%) |
| Walk-in centre, n/N (%) | 96/796 (12.1%) | 91/1204 (7.6%) | 1/56 (1.8%) | 95/740 (12.8%) | 27/157 (17.2%) | 68/583 (11.7%) |
| After recruitment in 28 day follow up period | | | | | | |
| Hospital admission/readmission | 63/795* (7.9%) | 78/1203 (6.5%) | 2/56 (3.6%) | 61/740 (8.2%) | 13/157 (8.3%) | 48/583 (8.2%) |

**Supplementary table 5 Number of healthcare contacts during illness (prior to recruitment and in 28-day follow up period)** PC = primary care; WIC = walk-in-centre; GP = General Practitioner; ED = Emergency Department *missing day 28 follow up data

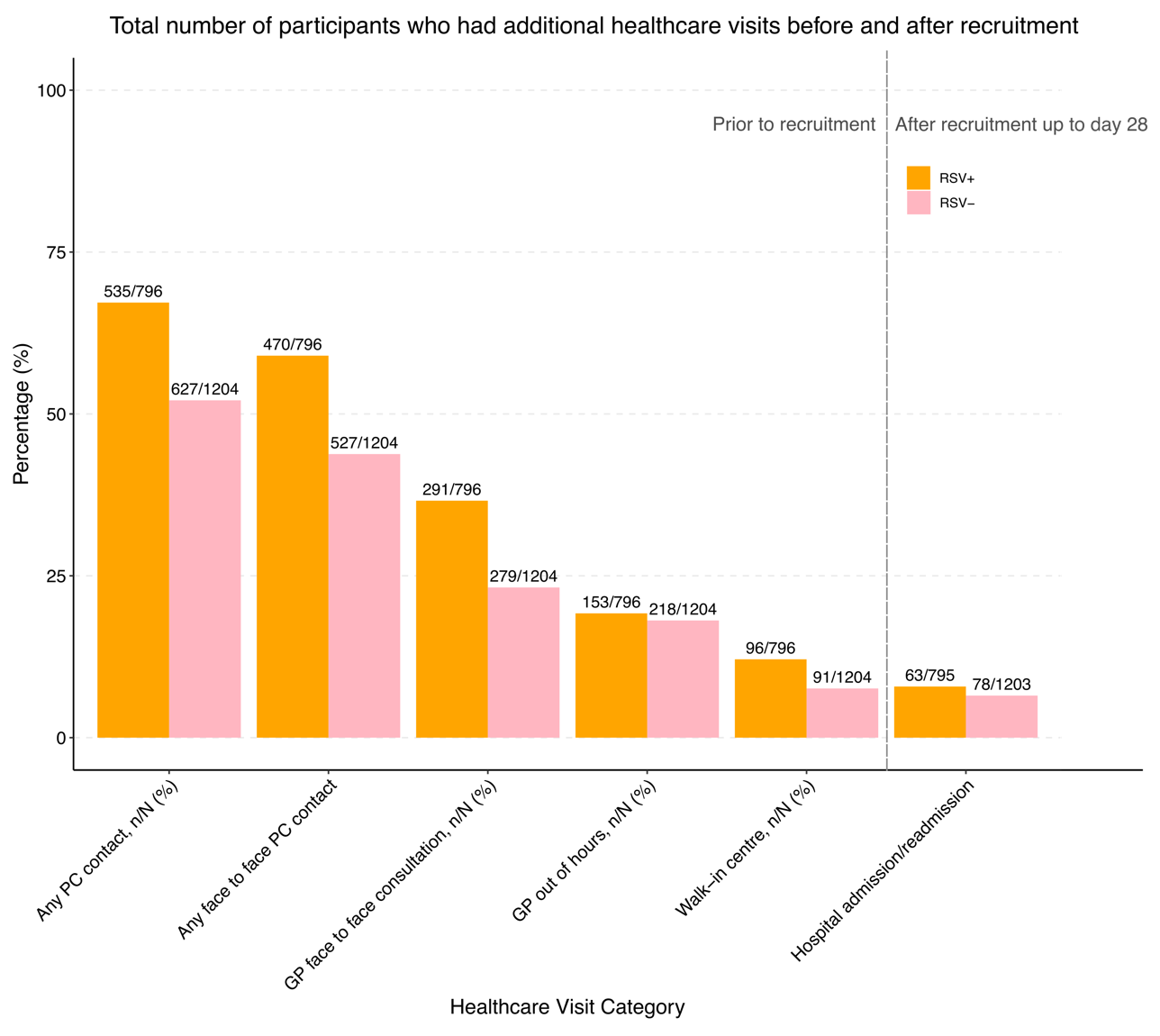

**Supplementary figure 4 Number of healthcare contacts during illness (prior to recruitment and in 28-day follow up period)** PC = primary care; WIC = walk-in-centre; GP = General Practitioner; ED = Emergency Department *missing day 28 follow up data

| **Multivariate analysis of risk factors for hospital admission in RSV positive participants** | | | | |
| --- | --- | --- | --- | --- |
|  | **Admitted to hospital 580/795 (73.0%)** | | | |
| **Risk factors** | **Risk factor not present n/N (%)** | **Risk factor present n/N (%)** | **Single Variable analysis** | **Multiple Variable analysis** |
| **Prematurity** |  |  |  |  |
| Any level of prematurity <37w (OR v not premature) | 482/682 (70.7%) | 98/113 (86.7%) | OR 2.71 (1.54, 4.78), p=0.001 | OR 2.25 (1.24, 4.06), p=0.007 |
| Extreme preterm <28w (OR v not premature) | 570/785 (72.6%) | 10/10 (100.0%) | * |  |
| Very preterm 28-31w (OR v not premature) | 565/779 (72.5%) | 15/16 (93.8%) | OR 6.22 (0.82, 47.43), p=0.078 |  |
| Mod-late preterm 32-36w (OR v not premature) | 507/708 (71.6%) | 73/87 (83.9%) | OR 2.16 (1.19, 3.92), p=0.011 |  |
| **Co-morbidity** |  |  |  |  |
| VW/asthma | 518/712 (72.8%) | 62/83 (74.7%) | OR 1.11 (0.66, 1.86), p=0.706 | OR 1.67 (0.95, 2.92), p=0.073 |
| BPD/CLD | 569/784 (72.6%) | 11/11 (100.0%) | * | * |
| Congenital cardiac disease | 557/768 (72.5%) | 23/27 (85.2%) | OR 2.18 (0.74, 6.37), p=0.155 | OR 2.21 (0.70, 6.94), p=0.175 |
| Other lung disease | 575/787 (73.1%) | 5/8 (62.5%) | OR 0.61 (0.15, 2.59), p=0.507 | OR 0.71 (0.16, 3.22), p=0.653 |
| Neurological | 572/787 (72.7%) | 8/8 (100.0%) | * | * |
| Other significant comorbidity | 562/774 (72.6%) | 18/21 (85.7%) | OR 2.26 (0.66, 7.76), p=0.194 | OR 1.86 (0.49, 6.97), p=0.360 |
| **Age** |  |  |  |  |
| Age <3m | 388/582 (66.7%) | 192/213 (90.1%) | OR 4.93 (2.67, 9.12), p<0.001 | OR 6.61 (3.45, 12.68), p<0.001 |
| Age 3-<6m | 460/633 (72.7%) | 120/162 (74.1%) | OR 1.54 (0.89, 2.66), p=0.120 | OR 1.98 (1.10, 3.55), p=0.022 |
| Age 6--<9m | 531/715 (74.3%) | 49/80 (61.3%) | OR 0.85 (0.46, 1.58), p=0.612 | OR 0.91 (0.48, 1.75), p=0.784 |
| Age 9-<12m | 531/724 (73.3%) | 49/71 (69.0%) | OR 1.20 (0.63, 2.31), p=0.581 | OR 1.37 (0.70, 2.70), p=0.361 |
| Age 12-<24m | 473/623 (75.9%) | 107/172 (62.2%) | OR 0.89 (0.53, 1.49), p=0.655 | OR 1.04 (0.61, 1.79), p=0.881 |
| Age 24-<36m | 517/698 (74.1%) | 63/97 (64.9%) | Ref | Ref |
| **Sex** |  |  |  |  |
| Sex (male) (OR v female) | 249/346 (72.5%) | 331/449 (73.7%) | OR 1.09 (0.80, 1.50), p=0.581 | OR 1.03 (0.74, 1.44), p=0.860 |
| **Season of birth** |  |  |  |  |
| Season of birth - Winter | 462/630 (73.3%) | 118/165 (71.5%) | Ref | Ref |
| Season of birth - Spring | 446/614 (72.6%) | 134/181 (74.0%) | OR 1.14 (0.71, 1.82), p=0.599 | OR 1.31 (0.79, 2.17), p=0.289 |
| Season of birth - Summer | 432/584 (74.0%) | 148/211 (70.1%) | OR 0.94 (0.60, 1.47), p=0.772 | OR 0.92 (0.57, 1.48), p=0.732 |
| Season of birth - Autumn | 401/558 (71.9%) | 179/237 (75.5%) | OR 1.23 (0.78, 1.93), p=0.368 | OR 1.00 (0.62, 1.63), p=0.990 |
| **Supplementary table 6 Multivariate analysis of risk factors for hospital admission in RSV positive participants**  Outcome of regression model (OR (95% CI), p-value) *All or zero exposed participants had outcome | | | | |
| Adjusted analyses are adjusted for age in months (continuous), sex, prematurity (<37 weeks), viral induced wheeze/asthma, bronchopulmonary dysplasia  HDU = high dependency unit, PICU = paediatric intensive care unit, VIW = viral induced wheeze, BPD = Bronchopulmonary dysplasia, CLD = chronic lung disease, OR = odds ratio; m = months; w = weeks | | | | |

| **Multivariate analysis of risk factors for hospital admission in RSV positive participants <12 months old** | | | | |
| --- | --- | --- | --- | --- |
|  | **Outcome of regression model (OR (95% CI), p-value)** | | | |
|  | **Admitted to hospital 410/526 (77.9%)** | | | |
| **Risk factors** | **Risk factor not present n/N (%)** | **Risk factor present n/N (%)** | **Single Variable** | **Multiple Variable** |
| **Prematurity** |  |  |  |  |
| **Any level of prematurity <37w (OR v not premature)** | 342/450 (76.0%) | 68/76 (89.5%) | OR 2.68 (1.25, 5.76), p=0.011 | OR 2.59 (1.17, 5.76), p=0.019 |
| **Extreme preterm <28w (OR v not premature)** | 405/521 (77.7%) | 5/5 (100.0%) | * |  |
| **Very preterm 28-31w (OR v not premature)** | 403/519 (77.6%) | 7/7 (100.0%) | * |  |
| **Mod-late preterm 32-36w (OR v not premature)** | 354/462 (76.6%) | 56/64 (87.5%) | OR 2.21 (1.02, 4.78), p=0.044 |  |
| **Co-morbidity** |  |  |  |  |
| **VW/asthma** | 392/500 (78.4%) | 18/26 (69.2%) | OR 0.62 (0.26, 1.46), p=0.276 | OR 0.80 (0.31, 2.08), p=0.649 |
| **BPD/CLD** | 407/523 (77.8%) | 3/3 (100.0%) | * | * |
| **Congenital cardiac disease** | 395/510 (77.5%) | 15/16 (93.8%) | OR 4.37 (0.57, 33.41), p=0.156 | OR 7.34 (0.90, 59.89), p=0.063 |
| **Other lung disease** | 407/522 (78.0%) | 3/4 (75.0%) | OR 0.85 (0.09, 8.23), p=0.887 | OR 1.00 (0.09, 10.69), p=0.999 |
| **Neurological** | 405/521 (77.7%) | 5/5 (100.0%) | * | * |
| **Other significant comorbidity** | 401/516 (77.7%) | 9/10 (90.0%) | OR 2.58 (0.32, 20.58), p=0.371 | OR 2.17 (0.24, 19.57), p=0.491 |
| **Sex** |  |  |  |  |
| **Sex (male)** | 170/223 (77.5%) | 240/303 (79.2%) | OR 1.19 (0.78, 1.80), p=0.416 | OR 1.15 (0.74, 1.80), p=0.527 |
| **Season of birth** |  |  |  |  |
| **Season of birth - Winter** | 329/418 (78.7%) | 80/107 (74.8%) | Ref | Ref |
| **Season of birth - Spring** | 313/406 (77.1%) | 96/119 (80.7%) | OR 1.41 (0.75, 2.65), p=0.287 | OR 1.84 (0.93, 3.65), p=0.079 |
| **Season of birth - Summer** | 304/384 (79.2%) | 105/141 (74.5%) | OR 0.98 (0.55, 1.75), p=0.957 | OR 0.99 (0.53, 1.85), p=0.972 |
| **Season of birth - Autumn** | 281/367 (76.6%) | 128/158 (81.0%) | OR 1.44 (0.80, 2.60), p=0.226 | OR 1.08 (0.57, 2.08), p=0.806 |
| **Supplementary table 7 Multivariate analysis of risk factors for hospital admission in RSV positive participants <12 months old** | | | | |
| Adjusted analyses are adjusted for age in months (continuous), sex, prematurity (<37 weeks), viral induced wheeze/asthma, bronchopulmonary dysplasia  HDU = high dependency unit, PICU = paediatric intensive care unit, VIW = viral induced wheeze, BPD = Bronchopulmonary dysplasia, CLD = chronic lung disease, OR = odds ratio; m = months; w = weeks. | | | | |

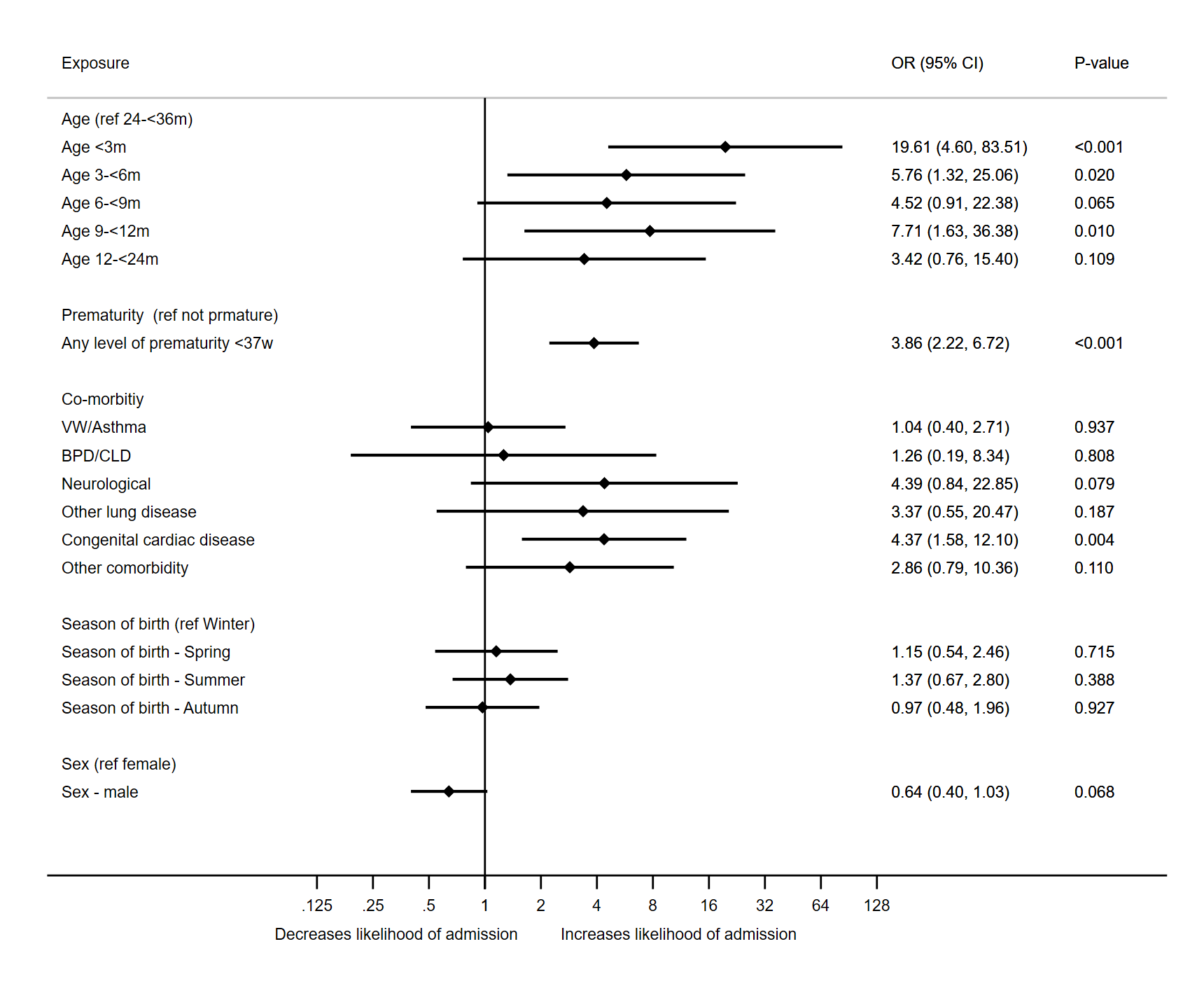

**Supplementary fig 5 Multivariable analysis of risk factors for admission to HDU or PICU in RSV-positive participants.** Adjusted analyses are adjusted for age in months (continuous), sex, prematurity (<37 weeks), viral induced wheeze/asthma, bronchopulmonary dysplasia. HDU = high dependency unit, PICU = paediatric intensive care unit, VIW = viral induced wheeze, BPD = Bronchopulmonary dysplasia, CLD = chronic lung disease, OR = odds ratio; m = months; w = weeks, RF=risk factor.

|  | **HDU or PICU admission by d28 90/795 (11.3%)** | | | |
| --- | --- | --- | --- | --- |
| **Risk factor** | **RF not present n/N (%)** | **RF present n/N (%)** | **Single Variable** | **Multiple Variable** |
| **Prematurity** |  |  |  |  |
| Any level of prematurity <37w (OR v not premature) | 60/682 (8.8%) | 30/113 (26.5%) | OR 3.75 (2.29, 6.14), p<0.001 | OR 3.86 (2.22, 6.72), p<0.001 |
| Extreme preterm <28w (OR v not premature) | 88/785 (11.2%) | 2/10 (20.0%) | OR 2.59 (0.54, 12.48), p=0.235 |  |
| Very preterm 28-31w (OR v not premature) | 86/779 (11.0%) | 4/16 (25.0%) | OR 3.46 (1.08, 11.05), p=0.037 |  |
| Mod-late preterm 32-36w (OR v not premature) | 66/708 (9.3%) | 24/87 (27.6%) | OR 3.95 (2.30, 6.77), p<0.001 |  |
| **Co-morbidity** |  |  |  |  |
| VW/asthma | 83/712 (11.7%) | 7/83 (8.4%) | OR 0.70 (0.31, 1.56), p=0.383 | OR 1.04 (0.40, 2.71), p=0.937 |
| BPD/CLD | 88/784 (11.2%) | 2/11 (18.2%) | OR 1.76 (0.37, 8.27), p=0.475 | OR 1.26 (0.19, 8.34), p=0.808 |
| Congenital cardiac disease | 81/768 (10.5%) | 9/27 (33.3%) | OR 4.24 (1.84, 9.75), p=0.001 | OR 4.37 (1.58, 12.10), p=0.004 |
| Other lung disease | 88/787 (11.2%) | 2/8 (25.0%) | OR 2.65 (0.53, 13.32), p=0.238 | OR 3.37 (0.55, 20.47), p=0.187 |
| Neurological | 86/787 (10.9%) | 4/8 (50.0%) | OR 8.15 (2.00, 33.18), p=0.003 | OR 4.39 (0.84, 22.85), p=0.079 |
| Other significant comorbidity | 85/774 (11.0%) | 5/21 (23.8%) | OR 2.53 (0.91, 7.09), p=0.077 | OR 2.86 (0.79, 10.36), p=0.110 |
| **Age** |  |  |  |  |
| Age <3m | 45/582 (7.7%) | 45/213 (21.1%) | OR 8.39 (2.54, 27.74), p<0.001 | OR 19.61 (4.60, 83.51), p<0.001 |
| Age 3-<6m | 74/633 (11.7%) | 16/162 (9.9%) | OR 3.43 (0.97, 12.11), p=0.055 | OR 5.76 (1.32, 25.06), p=0.020 |
| Age 6-<9m | 83/715 (11.6%) | 7/80 (8.8%) | OR 3.00 (0.75, 12.02), p=0.120 | OR 4.52 (0.91, 22.38), p=0.065 |
| Age 9-<12m | 81/724 (11.2%) | 9/71 (12.7%) | OR 4.55 (1.18, 17.46), p=0.027 | OR 7.71 (1.63, 36.38), p=0.010 |
| Age 12-<24m | 80/623 (12.8%) | 10/172 (5.8%) | OR 1.93 (0.52, 7.20), p=0.325 | OR 3.42 (0.76, 15.40), p=0.109 |
| Age 24-<36m | 87/698 (12.5%) | 3/97 (3.1%) | Ref | Ref |
| Sex |  |  |  |  |
| Sex (male) (OR v female) | 45/346 (10.5%) | 45/449 (10.0%) | OR 0.75 (0.48, 1.16), p=0.189 | OR 0.64 (0.40, 1.03), p=0.068 |
| **Season of birth** |  |  |  |  |
| Season of birth - Winter | 72/629 (11.4%) | 18/165 (10.9%) | Ref | Ref |
| Season of birth - Spring | 72/613 (11.7%) | 18/181 (9.9%) | OR 0.90 (0.45, 1.80), p=0.769 | OR 1.15 (0.54, 2.46), p=0.715 |
| Season of birth - Summer | 64/583 (11.0%) | 26/211 (12.3%) | OR 1.15 (0.61, 2.17), p=0.672 | OR 1.37 (0.67, 2.80), p=0.388 |
| Season of birth - Autumn | 62/557 (11.1%) | 28/237 (11.8%) | OR 1.09 (0.58, 2.05), p=0.779 | OR 0.97 (0.48, 1.96), p=0.927 |

**Supplementary table 8 risk factors for admission to HDU or PICU.** *All or zero exposed participants had outcome. Adjusted analyses are adjusted for age in months (continuous), sex, prematurity (<37 weeks), viral induced wheeze/asthma, bronchopulmonary dysplasia. HDU = high dependency unit, PICU = paediatric intensive care unit, VIW = viral induced wheeze, BPD = Bronchopulmonary dysplasia, CLD = chronic lung disease, OR = odds ratio; m = months; w = weeks, RF=risk factor

|  | **Oxygen/Ventilatory support** | | | | **Prescribed medication** | | |
| --- | --- | --- | --- | --- | --- | --- | --- |
|  | **High flow oxygen** | **CPAP** | **BIPAP** | **Intubation** | **Antibiotics** | **Inhalers** | **Steroids** |
| Overall | 158/760 (20.8%) | 17/759 (2.2%) | 12/760 (1.6%) | 35/760 (4.6%) | 189/796 (26.1%) | 157/796 (23.7%) | 55/796 (9.0%) |
| **Sex** |  |  |  |  |  |  |  |
| Male | 75/327 (22.9%) | 9/326 (2.8%) | 9/327 (2.8%) | 16/327 (4.9%) | 90/346 (28.6%) | 64/346 (22.2%) | 25/346 (9.4%) |
| Female | 83/433 (19.2%) (OR 0.80 (0.56, 1.13) p=0.206) | 8/433 (1.8%) (OR 0.66 (0.25, 1.74) p=0.403) | 3/433 (0.7%) (OR 0.25 (0.07, 0.92) p=0.037) | 19/433 (4.4%) (OR 0.89 (0.45, 1.76) p=0.742) | 99/450 (24.2%) (OR 0.80 (0.58, 1.11) p=0.188) | 93/450 (24.8%) (OR 1.15 (0.81, 1.64) p=0.446) | 30/450 (8.7%) (OR 0.92 (0.53, 1.59) p=0.758) |
| **Prematurity** | | | | | | | |
| Any level of prematurity (<37 weeks) | 40/112 (35.7%) (OR 2.50 (1.62, 3.85) p<0.001) | 4/112 (3.6%) (OR 1.81 (0.58, 5.64) p=0.309) | 5/112 (4.5%) (OR 4.28 (1.33, 13.73) p=0.015) | 13/112 (11.6%) (OR 3.74 (1.82, 7.66) p<0.001) | 33/113 (32.1%) (OR 1.39 (0.89, 2.17) p=0.142) | 21/113 (22.3%) (OR 0.92 (0.55, 1.53) p=0.742) | 10/113 (11.5%) (OR 1.38 (0.67, 2.82) p=0.382) |
| Extreme preterm <28w (OR v not premature) | 6/10 (60.0%) (OR 6.74 (1.87, 24.25) p=0.004) | 0/10 (0.0%) | 0/10 (0.0%) | 1/10 (10.0%) (OR 3.16 (0.38, 26.06) p=0.285) | 3/10 (33.0%) (OR 1.45 (0.37, 5.66) p=0.595) | 2/10 (24.0%) (OR 1.01 (0.21, 4.79) p=0.994) | 3/10 (39.0%) (OR 6.08 (1.52, 24.29) p=0.011) |
| Very preterm 28-31w (OR v not premature) | 7/16 (43.8%) (OR 3.49 (1.28, 9.57) p=0.015) | 1/16 (6.3%) (OR 3.25 (0.40, 26.48) p=0.271) | 1/16 (6.3%) (OR 6.10 (0.71, 52.77) p=0.100) | 1/16 (6.3%) (OR 1.90 (0.24, 15.01) p=0.544) | 8/16 (55.0%) (OR 3.38 (1.25, 9.15) p=0.017) | 6/16 (45.0%) (OR 2.41 (0.86, 6.76) p=0.093) | 1/16 (8.1%) (OR 0.95 (0.12, 7.32) p=0.957) |
| Mod-late preterm 32-36w (OR v not premature) | 27/86 (31.4%) (OR 2.06 (1.25, 3.38) p=0.005) | 3/86 (3.5%) (OR 1.76 (0.49, 6.32) p=0.384) | 4/86 (4.7%) (OR 4.47 (1.28, 15.59) p=0.019) | 11/86 (12.8%) (OR 4.17 (1.95, 8.94) p<0.001) | 22/87 (27.8%) (OR 1.14 (0.68, 1.91) p=0.610) | 13/87 (17.9%) (OR 0.71 (0.38, 1.31) p=0.271) | 6/87 (9.0%) (OR 1.05 (0.43, 2.54) p=0.913) |
| Not Premature (REF) | 118/648 (18.2%) | 13/647 (2.0%) | 7/648 (1.1%) | 22/648 (3.4%) | 156/683 (25.1%) | 136/683 (23.9%) | 45/683 (8.6%) |
| **Co-morbidity** | | | | | | | |
| Any significant co-morbidity | 34/131 (26.0%) (OR 1.43 (0.92, 2.21) p=0.111) | 3/131 (2.3%) (OR 1.03 (0.29, 3.63) p=0.966) | 4/131 (3.1%) (OR 2.44 (0.73, 8.24) p=0.149) | 8/131 (6.1%) (OR 1.45 (0.64, 3.27) p=0.370) | 47/133 (38.9%) (OR 2.01 (1.34, 2.99) p=0.001) | 53/133 (47.8%) (OR 3.56 (2.37, 5.34) p<0.001) | 23/133 (22.5%) (OR 4.12 (2.33, 7.31) p<0.001) |
| VIW/asthma | 17/83 (20.5%) (OR 0.98 (0.56, 1.72) p=0.942) | 1/83 (1.2%) (OR 0.50 (0.07, 3.84) p=0.508) | 1/83 (1.2%) (OR 0.74 (0.09, 5.79) p=0.773) | 1/83 (1.2%) (OR 0.23 (0.03, 1.71) p=0.151) | 21/83 (27.8%) (OR 1.10 (0.65, 1.86) p=0.725) | 45/83 (65.1%) (OR 6.35 (3.95, 10.23) p<0.001) | 17/83 (26.6%) (OR 4.58 (2.45, 8.55) p<0.001) |
| BPD/CLD | 5/11 (45.5%) (OR 3.25 (0.98, 10.78) p=0.054) | 0/11 (0.0%) | 0/11 (0.0%) | 2/11 (18.2%) (OR 4.82 (1.00, 23.21) p=0.050) | 6/11 (60.0%) (OR 3.95 (1.19, 13.08) p=0.025) | 4/11 (43.6%) (OR 2.36 (0.68, 8.17) p=0.175) | 4/11 (47.3%) (OR 8.22 (2.33, 29.02) p=0.001) |
| Other lung | 2/8 (25.0%) (OR 1.27 (0.25, 6.37) p=0.769) | 0/8 (0.0%) | 1/8 (12.5%) (OR 9.62 (1.09, 84.98) p=0.042) | 0/8 (0.0%) | 4/8 (55.0%) (OR 3.26 (0.81, 13.16) p=0.097) | 2/8 (30.0%) (OR 1.36 (0.27, 6.81) p=0.707) | 0/8 (0.0%) |
| Congenital cardiac disease | 13/26 (50.0%) (OR 4.06 (1.84, 8.95) p=0.001) | 2/26 (7.7%) (OR 3.99 (0.86, 18.43) p=0.076) | 1/26 (3.8%) (OR 2.63 (0.33, 21.16) p=0.364) | 4/26 (15.4%) (OR 4.12 (1.34, 12.69) p=0.014) | 11/27 (44.8%) (OR 2.28 (1.04, 5.01) p=0.040) | 2/27 (8.9%) (OR 0.32 (0.07, 1.35) p=0.121) | 5/27 (24.1%) (OR 3.27 (1.19, 9.00) p=0.022) |
| Neurological | 4/8 (50.0%) (OR 3.88 (0.96, 15.70) p=0.057) | 0/8 (0.0%) | 1/8 (12.5%) (OR 9.62 (1.09, 84.98) p=0.042) | 2/8 (25.0%) (OR 7.26 (1.41, 37.36) p=0.018) | 3/8 (41.3%) (OR 1.94 (0.46, 8.20) p=0.367) | 2/8 (30.0%) (OR 1.36 (0.27, 6.81) p=0.707) | 1/8 (16.3%) (OR 1.94 (0.23, 16.07) p=0.538) |
| No significant co-morbidity or prematurity (REF) | 98/551 (17.8%) | 10/550 (1.8%) | 6/551 (1.1%) | 16/551 (2.9%) | 121/584 (22.8%) | 94/584 (19.3%) | 27/584 (6.0%) |
| **Season of birth** | | | | | | | |
| Spring | 27/176 (15.3%) (OR 0.76 (0.43, 1.35) p=0.349) | 1/176 (0.6%) (OR 0.17 (0.02, 1.49) p=0.111) | 1/176 (0.6%) (OR 0.29 (0.03, 2.83) p=0.288) | 9/176 (5.1%) (OR 1.15 (0.42, 3.16) p=0.790) | 50/181 (30.4%) (OR 1.05 (0.65, 1.69) p=0.841) | 33/181 (21.9%) (OR 0.70 (0.41, 1.17) p=0.172) | 12/181 (8.6%) (OR 1.10 (0.46, 2.62) p=0.828) |
| Summer | 49/201 (24.4%) (OR 1.35 (0.81, 2.26) p=0.246) | 5/201 (2.5%) (OR 0.77 (0.22, 2.71) p=0.684) | 1/201 (0.5%) (OR 0.26 (0.03, 2.48) p=0.239) | 5/201 (2.5%) (OR 0.54 (0.17, 1.74) p=0.305) | 42/211 (21.9%) (OR 0.68 (0.42, 1.11) p=0.122) | 47/211 (26.7%) (OR 0.90 (0.55, 1.45) p=0.654) | 15/211 (9.2%) (OR 1.19 (0.52, 2.71) p=0.686) |
| Autumn | 51/226 (22.6%) (OR 1.22 (0.74, 2.03) p=0.433) | 6/225 (2.7%) (OR 0.83 (0.25, 2.76) p=0.758) | 7/226 (3.1%) (OR 1.63 (0.41, 6.40) p=0.484) | 14/226 (6.2%) (OR 1.41 (0.55, 3.57) p=0.474) | 52/238 (24.0%) (OR 0.77 (0.48, 1.22) p=0.265) | 37/238 (18.7%) (OR 0.58 (0.35, 0.95) p=0.030) | 18/238 (9.8%) (OR 1.27 (0.57, 2.82) p=0.560) |
| Winter (REF) | 30/156 (19.2%) | 5/156 (3.2%) | 3/156 (1.9%) | 7/156 (4.5%) | 44/165 (29.3%) | 40/165 (29.1%) | 10/165 (7.9%) |
| **Age** | | | | | | | |
| <3months (REF) | 65/213 (30.5%) (OR 6.22 (2.59, 14.97) p<0.001) | 12/213 (5.6%)* | 8/213 (3.8%)* | 22/213 (10.3%) (OR 10.37 (1.38, 78.12) p=0.023) | 46/214 (23.6%) (OR 0.64 (0.37, 1.11) p=0.110) | 4/214 (2.2%) (OR 0.02 (0.01, 0.06) p<0.001) | 5/214 (3.0%) (OR 0.08 (0.03, 0.22) p<0.001) |
| 3-<6months | 36/157 (22.9%) (OR 4.21 (1.70, 10.45) p=0.002) | 3/157 (1.9%)* | 2/157 (1.3%)* | 4/157 (2.5%) (OR 2.35 (0.26, 21.38) p=0.447) | 27/162 (18.3%) (OR 0.47 (0.26, 0.85) p=0.013) | 7/162 (5.2%) (OR 0.05 (0.02, 0.11) p<0.001) | 4/162 (3.2%) (OR 0.09 (0.03, 0.26) p<0.001) |
| 6-<9months | 13/74 (17.6%) (OR 3.02 (1.09, 8.39) p=0.034) | 0/74 (0.0%)* | 0/74 (0.0%)* | 3/74 (4.1%) (OR 3.80 (0.39, 37.34) p=0.252) | 14/80 (19.3%) (OR 0.50 (0.24, 1.02) p=0.058) | 2/80 (3.0%) (OR 0.03 (0.01, 0.11) p<0.001) | 1/80 (1.6%) (OR 0.04 (0.01, 0.33) p=0.002) |
| 9-<12 months | 17/66 (25.8%) (OR 4.91 (1.82, 13.29) p=0.002) | 1/65 (1.5%)* | 1/66 (1.5%)* | 2/66 (3.0%) (OR 2.81 (0.25, 31.68) p=0.403) | 19/71 (29.4%) (OR 0.86 (0.43, 1.69) p=0.657) | 15/71 (25.4%) (OR 0.27 (0.14, 0.55) p<0.001) | 3/71 (5.5%) (OR 0.15 (0.04, 0.53) p=0.003) |
| 12-<24 months | 21/159 (13.2%) (OR 2.16 (0.84, 5.56) p=0.112) | 1/159 (0.6%)* | 1/159 (0.6%)* | 3/159 (1.9%) (OR 1.73 (0.18, 16.89) p=0.637) | 54/172 (34.5%) (OR 1.07 (0.62, 1.84) p=0.798) | 81/172 (56.5%) (OR 0.91 (0.55, 1.50) p=0.706) | 20/172 (15.1%) (OR 0.45 (0.23, 0.87) p=0.018) |
| 24-<36 months | 6/91 (6.6%) | 0/91 (0.0%) | 0/91 (0.0%) | 1/91 (1.1%) | 29/97 (32.9%) | 48/97 (59.4%) | 22/97 (29.5%) |

**Supplementary table 9 Odds ratios for medical interventions by age, sex, prematurity, co-morbidity status, season of birth.**
HDU = high dependency unit, PICU = paediatric intensive care unit, VIW = viral induced wheeze, BPD = Bronchopulmonary dysplasia, CLD = chronic lung disease, OR=odds ratio, CPAP = continuous positive airway pressure, BiPAP = Biphasic positive airway pressure

*No OR as no events in reference population

| **RSV positive** | **Overall medically attended** | **Hospitalised** | **Not hospitalised** | **HDU** | **PICU** |
| --- | --- | --- | --- | --- | --- |
| **Overall** | 795* | 580/795 (73.0%) | 215/795 (27.0%) | 60/795 (7.5%) | 38/60 (63.3%) |
| <3m | 213/795 (26.7%) | 192/580 (33.1%) | 21/215 (9.8%) | 27/60 (45.0%) | 22/38 (57.9%) |
| 3-<6m | 162/795 (20.3%) | 120/580 (20.7%) | 42/215 (19.5%) | 11/60 (18.3%) | 5/38 (13.2%) |
| 6-<9m | 80/795 (10.1%) | 49/580 (8.4%) | 31/215 (14.4%) | 5/60 (8.3%) | 3/38 (7.9%) |
| 9-<12m | 71/795 (8.9%) | 49/580 (8.4%) | 22/215 (10.2%) | 7/60 (11.7%) | 3/38 (7.9%) |
| **Total Age <12m** | 526/795 (66.2%) | 410/580 (70.7%) | 116/215 (54.0%) | 50/60 (83.3%) | 33/38 (86.8%) |
| **Age 12-<24m** | 172/795 (21.4%) | 107/580 (18.4%) | 65/215 (30.2%) | 9/60 (15.0%) | 3/38 (7.9%) |
| **Age 24m-<36m** | 97/795 (12.2%) | 63/580 (10.9%) | 34/215 (15.8%) | 1/60 (1.7%) | 2/38 (5.3%) |
| **Total with Significant underlying co-morbidity and/or prematurity**  **Total without significant underlying co-morbidity or prematurity** | 212/795 (26.7%)  583/795 (73.3%) | 169/580 (29.1%)  411/580 (70.1%) | 43/215 (20.0%)  172/215 (80.0%) | 27/60 (45.0%)  33/60 (55%) | 22/38 (57.9%)  16/38 (42.1%) |

**Supplementary table 10** Proportion of hospitalised, HDU, PICU RSV positive participants in each age category and with co-morbidity/prematurity

HDU = high dependency unit, PICU = paediatric intensive care unit, m = months, w = weeks, ***** 1 missing day 28 follow up

|  | **Prolonged respiratory symptoms at d14 335/418 (80.1%)** | | | |
| --- | --- | --- | --- | --- |
| **Risk factor** | **No n/N (%)** | **Yes n/N (%)** | **Single Variable** | **Multiple Variable** |
| **Prematurity** |  |  |  |  |
| Any level of prematurity <37w (OR v not premature) | 291/364 (79.9%) | 44/54 (81.5%) | OR 1.10 (0.53, 2.30), p=0.792 | OR 1.26 (0.58, 2.76), p=0.559 |
| Extreme preterm <28w (OR v not premature) | 332/414 (80.2%) | 3/4 (75.0%) | OR 0.75 (0.08, 7.34), p=0.807 |  |
| Very preterm 28-31w (OR v not premature) | 329/411 (80.0%) | 6/7 (85.7%) | OR 1.51 (0.18, 12.70), p=0.707 |  |
| Mod-late preterm 32-36w (OR v not premature) | 300/375 (80.0%) | 35/43 (81.4%) | OR 1.10 (0.49, 2.47), p=0.822 |  |
| **Co-morbidity** |  |  |  |  |
| VW/asthma | 302/376 (80.3%) | 33/42 (78.6%) | OR 0.90 (0.41, 1.96), p=0.788 | OR 0.87 (0.37, 2.02), p=0.744 |
| BPD/CLD | 333/415 (80.2%) | 2/3 (66.7%) | OR 0.49 (0.04, 5.50), p=0.565 | OR 0.61 (0.02, 15.21), p=0.766 |
| Congenital cardiac disease | 324/405 (80.0%) | 11/13 (84.6%) | OR 1.38 (0.30, 6.33), p=0.683 | OR 1.23 (0.23, 6.68), p=0.812 |
| Other lung disease | 331/413 (80.1%) | 4/5 (80.0%) | OR 0.99 (0.11, 8.98), p=0.994 | OR 1.00 (0.10, 9.50), p=0.999 |
| Neurological | 334/416 (80.3%) | 1/2 (50.0%) | OR 0.25 (0.02, 3.97), p=0.323 | OR 0.22 (0.01, 7.05), p=0.394 |
| Other significant comorbidity | 326/409 (79.7%) | 9/9 (100.0%) | * | * |
| **Age** |  |  |  |  |
| Age <3m | 246/309 (99.2%) | 89/109 (81.7%) | OR 1.40 (0.61, 3.22), p=0.430 | OR 1.27 (0.53, 3.09), p=0.592 |
| Age 3-5m | 273/338 (99.2%) | 62/80 (77.5%) | OR 1.08 (0.46, 2.55), p=0.856 | OR 1.08 (0.43, 2.68), p=0.876 |
| Age 6-8m | 299/374 (99.2%) | 36/44 (81.8%) | OR 1.41 (0.51, 3.93), p=0.506 | OR 1.46 (0.51, 4.22), p=0.485 |
| Age 9-11m | 300/376 (99.2%) | 35/42 (83.3%) | OR 1.57 (0.55, 4.52), p=0.402 | OR 1.53 (0.52, 4.49), p=0.440 |
| Age 12-23m | 257/321 (99.2%) | 78/97 (80.4%) | OR 1.29 (0.56, 3.00), p=0.553 | OR 1.27 (0.54, 3.01), p=0.581 |
| Age 24-35m | 300/372 (99.2%) | 35/46 (76.1%) | Ref | Ref |
| **Sex** |  |  |  |  |
| Sex (male) | 153/189 (80.0%) | 182/229 (79.5%) | OR 0.91 (0.56, 1.48), p=0.707 | OR 0.94 (0.57, 1.54), p=0.805 |
| **Season of birth** |  |  |  |  |
| Season of birth - Winter | 265/325 (80.0%) | 70/93 (75.3%) | Ref | Ref |
| Season of birth - Spring | 257/321 (80.0%) | 78/97 (80.4%) | OR 1.35 (0.68, 2.68), p=0.394 | OR 1.30 (0.65, 2.63), p=0.459 |
| Season of birth - Summer | 254/315 (80.0%) | 81/103 (78.6%) | OR 1.21 (0.62, 2.36), p=0.575 | OR 1.33 (0.67, 2.62), p=0.418 |
| Season of birth - Autumn | 229/293 (80.0%) | 106/125 (84.8%) | OR 1.83 (0.93, 3.61), p=0.080 | OR 1.83 (0.91, 3.67), p=0.090 |

**Supplementary table 11 multivariate analysis of risk factors for prolonged respiratory symptoms d14 Outcome of regression model (OR (95% CI), p-value)**

*All or zero exposed participants had outcome. Adjusted analyses are adjusted for age in months (continuous), sex, prematurity (<37 weeks), viral induced wheeze/asthma, bronchopulmonary dysplasia. HDU = high dependency unit, PICU = paediatric intensive care unit, VIW = viral induced wheeze, BPD = Bronchopulmonary dysplasia, CLD = chronic lung disease, OR = odds ratio; m = months; w = weeks

|  |  | **Total RSV +ve with coinfection (RSV + with 1 or more additional virus)** | **RSV +ve coinfection with 2 virus (RSV + 1 other)** | **RSV +ve coinfection with 3 or more virus (RSV + 2 or more others)** | **Total RSV –ve with coinfection (2 or more viruses)** | **RSV -ve coinfection with 2 viruses (2 viruses not RSV)** | **RSV –ve infection with 2 or more virus (3 or more viruses not RSV)** |
| --- | --- | --- | --- | --- | --- | --- | --- |
| **Overall** | | 325/796 (40.8%) | 255/796 (32.0%) | 66/796 (8.3%) | 253/1204 (21.0%) | 220/1204 (18.3%) | 30/1204 (2.5%) |
| **Age** | |  |  |  |  |  |  |
|  | <3m | 59/214 (27.6%) | 51/214 (23.8%) | 6/214 (2.8%) | 21/150 (14.0%) | 18/150 (12.0%) | 2/150 (1.3%) |
|  | 3-<6m | 47/162 (29.0%) | 41/162 (25.3%) | 6/162 (3.7%) | 25/132 (18.9%) | 22/132 (16.7%) | 3/132 (2.3%) |
|  | 6-<9m | 32/80 (40.0%) | 25/80 (31.3%) | 7/80 (8.8%) | 28/129 (21.7%) | 26/129 (20.2%) | 2/129 (1.6%) |
|  | 9-<12m | 46/71 (64.8%) | 34/71 (47.9%) | 11/71 (15.5%) | 35/136 (25.7%) | 30/136 (22.1%) | 4/136 (2.9%) |
|  | 12-<24m | 89/172 (51.7%) | 70/172 (40.7%) | 19/172 (11.0%) | 99/405 (24.4%) | 86/405 (21.2%) | 13/405 (3.2%) |
|  | 24-<36m | 52/97 (53.6%) | 34/97 (35.1%) | 17/97 (17.5%) | 45/252 (17.9%) | 38/252 (15.1%) | 6/252 (2.4%) |
| **Season of recruitment** | |  |  |  |  |  |  |
|  | Spring | 65/181 (35.9%) | 55/181 (30.4%) | 10/181 (5.5%) | 62/259 (23.9%) | 53/259 (20.5%) | 7/259 (2.7%) |
|  | Summer | 91/211 (43.1%) | 70/211 (33.2%) | 19/211 (9.0%) | 61/292 (20.9%) | 54/292 (18.5%) | 7/292 (2.4%) |
|  | Autumn | 89/238 (37.4%) | 66/238 (27.7%) | 22/238 (9.2%) | 53/294 (18.0%) | 44/294 (15.0%) | 8/294 (2.7%) |
|  | Winter | 80/165 (48.5%) | 64/165 (38.8%) | 15/165 (9.1%) | 77/358 (21.5%) | 69/358 (19.3%) | 8/358 (2.2%) |
| **Site of recruitment** | |  |  |  |  |  |  |
|  | Primary care | 21/51 (41.2%) | 15/51 (29.4%) | 6/51 (11.8%) | 43/232 (18.5%) | 41/232 (17.7%) | 1/232 (0.4%) |
|  | WIC | 3/5 (60.0%) | 3/5 (60.0%) | 0/5 (0.0%) | 19/122 (15.6%) | 16/122 (13.1%) | 3/122 (2.5%) |
|  | ED+discharge | 73/157 (46.5%) | 56/157 (35.7%) | 16/157 (10.2%) | 53/276 (19.2%) | 47/276 (17.0%) | 5/276 (1.8%) |
|  | Hospital admission (direct or via ED+admission) | 228/583 (39.1%) | 181/583 (31.0%) | 44/583 (7.5%) | 138/574 (24.0%) | 116/574 (20.2%) | 21/574 (3.7%) |

**Supplementary table 12 Rates of Viral Coinfection by age group, season and site of recruitment**

ED = emergency department, WIC = walk in centre, m = months

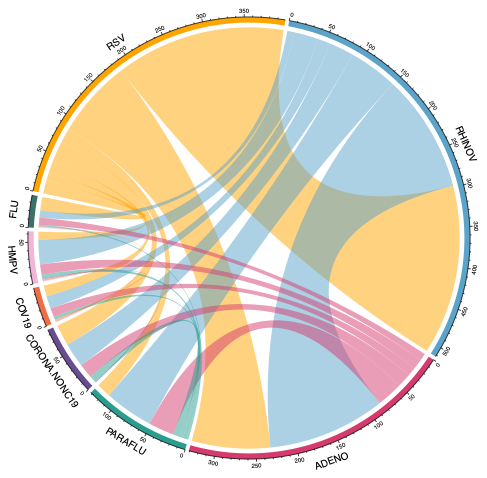

Supplementary figure 6

Chord diagram demonstrating proportion of each virus involved in co-infection

RSV = Respiratory Syncytial virus, RHINOV=rhinovirus, ADENO=adenovirus, CORNA-NONc19= non-SARS-CoV-2 coronavirus, PARAFLU=parainfluenza virus, COV—19- SARS-CoV-2, HMPV=human metapneumovirus

**HE analysis**

**Supplementary Table. Cost table**

| **Description** | **Unit cost** |
| --- | --- |
| GP telephone consultation | £11.40 |
| GP consultation | £36.00 |
| GP out of hours | £93.70 |
| Walk-in centre | £93.70 |
| A&E attendance | £185.00 |
| Ward admission – Short (<3 days) | £1100.23 |
| Ward admission – Short & Long (≥ 3 days) | £1909.86 |
| High Dependency / Paediatric Intensive Care Unit | £2905.20 |

**Supplementary Table Healthcare costs associated with the treatment of RSV**

| **RSV status** | **N** | **Mean (SD)** | **Min; Max** | **95%CI** | **Total costs** |
| --- | --- | --- | --- | --- | --- |
| Negative | 949 | £1294.58 (£1179.39) | £11.4 ; £5210.26 | £1219.54 ; £1369.62 | £1,228,554.76 |
| Positive | 747 | £1811.44 (£1281.25) | £11.4 ; £5187.46 | £1719.56 ; £1903.32 | £1,353,144.43 |
| Overall |  | £1522.23 (£1251.53) |  | £1462.66 ; £1581.79 | £2,581,699.19 |
